# Standardized and optimized preservation, extraction and quantification techniques for detection of fecal SARS-CoV-2 RNA

**DOI:** 10.1101/2021.04.10.21255250

**Authors:** Aravind Natarajan, Alvin Han, Soumaya Zlitni, Erin F. Brooks, Summer E. Vance, Marlene Wolfe, Upinder Singh, Prasanna Jagannathan, Benjamin A. Pinsky, Alexandria Boehm, Ami S. Bhatt

## Abstract

COVID-19 patients shed SARS-CoV-2 viral RNA in their stool, sometimes well after they have cleared their respiratory infection. This feature of the disease may be significant for patient health, epidemiology, and diagnosis. However, to date, methods to preserve stool samples from COVID patients, and to extract and quantify viral RNA concentration have yet to be optimized. We sought to meet this urgent need by developing and benchmarking a standardized protocol for the fecal detection of SARS-CoV-2 RNA. We test three preservative conditions for their ability to yield detectable SARS-CoV-2 RNA: OMNIgene-GUT, Zymo DNA/RNA shield kit, and the most common condition, storage without any preservative. We test these in combination with three extraction kits: the QIAamp Viral RNA Mini Kit, Zymo Quick-RNA Viral Kit, and MagMAX Viral/Pathogen Kit. Finally, we also test the utility of two detection methods, ddPCR and RT-qPCR, for the robust quantification of SARS-CoV-2 viral RNA from stool. We identify that the Zymo DNA/RNA shield collection kit and the QiaAMP viral RNA mini kit yield more detectable RNA than the others, using both ddPCR and RT-qPCR assays. We also demonstrate key features of experimental design including the incorporation of appropriate controls and data analysis, and apply these techniques to effectively extract viral RNA from fecal samples acquired from COVID-19 outpatients enrolled in a clinical trial. **Finally, we** recommend a comprehensive methodology for future preservation, extraction and detection of RNA from SARS-CoV-2 and other coronaviruses in stool.

## Introduction

Severe acute respiratory syndrome coronavirus-2 (SARS-CoV-2) is an RNA virus from the *Coronaviridae* family^1^ that causes coronavirus disease 2019 (COVID-19). This disease has spread rapidly across the globe and remains a public health threat ^2^. COVID-19 is typically considered a respiratory disease, with primary symptoms including cough, sore throat, congestion, anosmia, and dyspnea. However, gastrointestinal (GI) symptoms are also recognized as manifestations of the disease^3, 4^. Further, patients are found to shed viral RNA in their stool up to even 4 months after disease onset, well after they have cleared the infection from their respiratory tissues (unpublished observation, manuscript in preparation). While transmission of SARS-CoV-2 typically occurs through the respiratory tract, some reports indicate the presence of infectious viral particles in patient stool^5, 6^. Whether these are truly infectious and have ramifications for public health remains to be definitively demonstrated. However, from an individual patient health perspective, SARS-CoV-2 antigen is found to persist in the GI tract, promoting evolution of host humoral immunity to variants of the virus^7^. Relatedly, prolonged viral RNA shedding in stool may indicate a superior immune response^7^. Finally, from an epidemiological perspective, researchers monitor SARS-CoV-2 load in sewage as a proxy for the burden of disease within a population^8^. Taken together, monitoring the fecal shedding of SARS-CoV-2 is vital to fully understanding this pathogen and its effect on patient health in addition to informing public health measures. Therefore, a standardized method to handle and process samples for accurate quantification of viral RNA in stool is critical. Notably, the proposed method should allow for external validity and harmonization of data across studies.

Accurately quantifying fecal shedding of SARS-CoV-2 RNA is challenging. Stool is a heterogeneous sample matrix that contains numerous PCR inhibitors that impede downstream processes like reverse-transcriptase quantitative polymerase chain reaction (RT-qPCR) for quantifying RNA ^9^. Further, stool also contains RNases that can rapidly degrade unprotected RNA. Therefore, it is critical that we use appropriate preservatives that protect RNA in stool and employ extraction methods that effectively recover RNA without co-eluting inhibitors or contaminants. In the absence of a comprehensive, standardized protocol, existing studies of SARS-CoV-2 RNA in stool employ methods that have not yet been optimized. Further, the variability in techniques used across studies makes meta-analysis difficult, hindering our overall understanding of the disease. While there is heterogeneity, the majority of existing studies collect and store stool without any preservative^10–12^, dilute stool in PBS at the time of RNA extraction, and employ the QIA-Amp Viral RNA kit from Qiagen for RNA isolation. Unfortunately, the efficiency of these strategies in preserving and extracting SARS-CoV-2 RNA is unknown and has not yet been systematically analyzed. Finally, after RNA extraction, the detection and quantification of RNA by RT-qPCR has elements that have yet to be standardized. While the primer/probe sets that are used are generally consistent, the calling of “positivity” for SARS-CoV-2 RNA often relies on arbitrary thresholds set in the absence of a relevant standard curve^13–15^. These experimental inconsistencies and the lack of a clearly validated experimental pipeline contribute significantly to heterogeneity in detection and quantification of viral RNA in stool. To overcome these challenges, we sought to test a variety of accessible and common methods for the preservation of stool samples, RNA extraction and detection of viral RNA from stool samples, and present here an optimized pipeline.

Therefore, in the current study, we present data comparing the performance of three different stool preservatives, three nucleic acid extraction kits, and two PCR based assays for detecting fecal SARS-CoV-2 RNA. Based on these data, we recommend a pipeline for collecting and processing stool samples for the detection of SARS-CoV-2 RNA. Finally, we validate this standardized pipeline using patient samples collected from a clinical trial. Altogether, our findings here will guide the field towards a more standardized method of robustly measuring the fecal burden of SARS-CoV-2 RNA both in clinical and research settings.

## Methods

### Preparation of stool samples spiked with SARS-CoV-2 RNA or BCoV attenuated virus

We used two types of non-clinical stool samples in this study. The first set of samples were acquired from the United States National Institute of Standards and Technology (NIST)^16^ and stored at −80°C. The second were acquired from healthy donors collected as part of the Stanford IRB protocol #42043 (PI: Ami S Bhatt; Title : Genomic, Transcriptomic and Microbiological Characterization of Human Body Fluid Specimens) and stored at −80°C without any preservatives.

In most studies, stool samples are collected and stored without a preservative^10–12^. They are then resuspended in PBS in a 1:5 ratio (w:v) prior to RNA extraction. As a proxy for these samples, we added 1000 mg of stool to 5 ml of PBS, and describe these as PBS preserved samples through this study. Separately, we also stored samples in the OMNIgene-GUT tube (OG; DNA Genotek; Catalog # OMR-200) and Zymo DNA/RNA Shield (ZY; Zymo Research; Catalog # R1100-250) kit according to the manufacturer’s instructions. Briefly, we added 500 mg of stool to the OG tube containing 2 mL of preservative to prepare the OG samples, and 1000 mg stool to the ZY kit with 9 mL of buffer to prepare the ZY samples. Given shortages in the supply of the ZY kit, we also resorted to recreating this kit in house using 9 mL of the DNA/RNA Shield buffer (Zymo Research; Catalog # R1100-1L; Lot # ZRC195881) in a 15 mL centrifuge tube (VWR; Catalog # 89039-666). Subsequently, where listed, we spiked in synthetic SARS-CoV-2 RNA from ATCC (Catalog # VR-3276SD, Lot # 70034237) at a final concentration of either 10^3^ or 10^4^ copies per µL of preserved stool sample. For samples spiked with the control bovine coronavirus (BCoV), we prepared BCoV by resuspending one vial of lyophilized Zoetis Calf-Guard Bovine Rotavirus-Coronavirus Vaccine (Catalog # VLN 190/PCN 1931.20) in 3 mL of PBS to create an undiluted reagent as per manufacturer’s instructions. We then added 60 µL of either undiluted or diluted BCoV (1:10 in PBS) to 3 mL of preserved stool sample. To create an extraction blank control, an equivalent volume of PBS was added to samples labelled “None”. All samples were then stored in 1.5 mL DNA LoBind tubes (Fisher Scientific; Catalog # 13-698-791) and immediately frozen at −80°C.

### Preparation of clinical stool samples

Clinical samples were collected and stored from patients participating in an interventional study of Peginterferon Lambda 1a^17^ as described in the original study (manuscript in preparation). Briefly, study subjects were requested to collect samples in both the OG and ZY tubes according to manufacturer’s instructions, and samples were stored at room temperature for up to seven days before being processed into cryovials and frozen at -80C until subsequent use. We directly used OG and ZY preserved samples in the subsequent extraction steps. Where mentioned samples were spiked with 10 μL of attenuated BCoV vaccine per 500 μL of preserved stool sample after thawing an aliquot for extraction.

### Viral RNA extraction

We spun down 600 µL of each preserved stool sample at 10,000x *g* for 2 minutes to remove solids from the sample. We then processed 200 µL of supernatant according to manufacturer’s instructions for the QIAamp Viral RNA Mini Kit (Qiagen; Catalog # 52906, Lot #166024216) and Zymo Quick-RNA Viral Kit (Zymo; Catalog # R1035, Lot #206187). In the supernatant processed using the MagMAX Viral/Pathogen Kit (Thermofisher Scientific; Catalog # A42352, Lot #’s 2009063, 2008058) we followed the manufacturer’s instructions with the following small exception: samples were processed in 1.5 mL DNA LoBind tubes rather than 1.5 mL deep well plates. We eluted RNA from each sample in 60 µL of the elution buffer included in each kit. The eluted RNA was stored in a 96-well plate at −80°C.

### Quantification of viral RNA by RT-qPCR

We assembled the RT-qPCR reaction using a Biomek FX liquid handler, adding 5 µL of eluted RNA to 5 µL of TaqPath 1-Step RT-qPCR CG mastermix (Applied Biosystems, Catalog # A15300, Lot 2293196), 8.5 µL of nuclease-free water (Ambion, Catalog # AM9937, Lot 2009117), and 1.5 uL of primer/probe mix. The primer/probe mix was composed of 200 nM each of forward primer, reverse primer and probe (Elim Biopharmaceuticals) with sequences summarized in Table 1. We designed the probes to bear a 5’ Fluorescein (FAM) and 3’ 5- Carboxytetramethylrhodamine (TAMRA) dyes.

**Table 1.** Sequences of oligonucleotides used as primers and probes in this study.

| **Sample ID** | **Wet weight** | **Dry weight** | **Percent stool (%)** | **Wet weight** | **Dry weight** | **Percent stool** |
| --- | --- | --- | --- | --- | --- | --- |
| Lambda_248 | 0.119 | 0.036 | 30.252 | 0.172 | 0.031 | 18.023 |
| Lambda_327 | 0.145 | 0.043 | 29.655 | 0.117 | 0.011 | 9.402 |
| Lambda_223 | 0.101 | 0.034 | 33.663 | 0.168 | 0.023 | 13.69 |
| Lambda_264 | 0.096 | 0.031 | 32.292 | 0.117 | 0.018 | 15.385 |
| Lambda_292 | 0.157 | 0.049 | 31.21 | 0.111 | 0.013 | 11.712 |
| **Average** |  |  | 31.414 |  |  | 13.642 |
| **Std deviation** |  |  | 1.606 |  |  | 3.314 |

Our RT-qPCR analysis is guided by the Minimum Information for Publication of Quantitative Real-Time PCR Experiments (MIQE) guidelines^18^. We used the QuantStudio 12K Flex (Applied Biosystems) to amplify the template using the following thermocycling program: 25°C for 2 min, 50°C for 15 min, 95°C for 2 min, 45 cycles of 95°C for 15 sec and 55°C for 30 sec with ramp speed of 1.6°C per second at each step. We calculated the quantification cycle (C_q_) value using the system software. Standard curves for quantification were generated using a five-point ten-fold dilution of the SARS-CoV-2 ATCC standard from 10^4^ to 10^0^ copies per µL of template. We calculated the concentration of RNA using a linear regression of the standard curve. We established limit of blank on a plate-by-plate basis; specifically, we turned to the specific plate that an experimental sample was assayed on and picked the lowest C_q_ among the following controls run in the same plate: the y-intercept of the line of best fit from the standard curve, none (no RNA or BCoV spiked) stool samples, water and elution buffers from the RNA extraction kits as listed in the relevant experiments. RNA concentrations from reactions with C_q_ values below the LoB were defined as “undetermined”. The concentration of RNA from technical duplicate RT-qPCR reactions were averaged. If one of the two technical duplicate reactions failed to amplify within the range of the standard curve, the viral concentration from that sample was treated as ‘Undetermined’.

### Quantification of viral RNA by ddPCR

Our ddPCR analysis is guided by the Droplet Digital PCR Applications Guide on QX200 machines (BioRad)^19^. We assembled the ddPCR reaction using a Biomek FX liquid handler, by adding 5.5 µL of eluted RNA to 5.5 µL Supermix, 2.2 µL reverse transcriptase, 1.1 µL of 300 nM Dithiothreitol (DTT), 1.1 µL of 20x Custom ddPCR Assay Primer/Probe Mix (BioRad, Catalog # 10031277) and 6.6 µL of nuclease-free water (Ambion, Catalog # AM9937, Lot 2009117). The Supermix, reverse transcriptase and DTT are from the One-Step RT-ddPCR Advanced Kit for Probes (BioRad, Catalog # 1864021). We then processed the assembled reactions on a QX200 AutoDG Droplet Digital PCR System to ensure consistency in droplet generation across samples. Amplification was performed on a BioRad T100 thermocycler using the following thermocycling program: 50°C for 60 min, 95°C for 10 min, 40 cycles of 94°C for 30 sec and 55°C for 1 min, followed by 1 cycle of 98°C for 10 min and 4°C for 30 min with ramp speed of 1.6°C per second at each step^20^.

We thresholded the samples to ascertain the value at which a droplet was considered “positive” by applying a multistep process that used the following positive and negative controls included on each plate: ATCC SARS-CoV-2 RNA, RNA extracted directly from attenuated BCoV vaccine prepared in PBS, water and elution buffers. First, we set the threshold between the mean positive and negative amplitudes of these controls to minimize detected copies in the negative controls and to reflect the expected RNA concentration of the positive samples. We then calculated the difference between the mean negative amplitude and the threshold amplitude in the negative control reactions and added it to the mean negative amplitude for each sample.

Further, we noted the highest detected copy number in the none (no RNA or BCoV spiked) stool samples as the LoB. Samples with detected copies per µL below the LoB were marked as “Undetermined”. Finally, absolute quantification of nucleic acids using ddPCR relies on the generation of a Poisson distribution of template RNA in droplets, requiring an adequate number of droplets with a negative amplification signal. Therefore, in instances where a reaction has saturated amounts of template, we diluted the sample and performed the assay again to ensure reliable quantification. These dilutions are listed where they were performed. Final copy numbers are reported as copies per µL of target in eluate. This was calculated by multiplying by copies per µL reported in each ddPCR reaction by total reaction volume (22 µL) and dividing by input template volume (5.5 µL).

### Measurement of dry weight

We first noted down the weight of one 1.5 mL DNA LoBind microcentrifuge tube per sample. Next, we took two biopsy punches using the Integra Miltex Biopsy Punches with Plunger System (Thermo Fisher Scientific; Catalog # 12-460-410) from each of the relevant stool samples and transferred these to microcentrifuge tubes corresponding to the respective samples. The tubes were then weighed, and the respective wet weight was calculated upon subtracting the weight of the empty tube. Next, they were incubated on a heat block at 100°C for 72 hours and reweighed. The dry weight was calculated upon subtracting the weight of the empty tube.

### Data analysis and statistics

We performed statistical analyses using R (version 4.0.0). All statistical analyses were two-sided, performed on the data prior to rounding and statistical significance was assessed at α = 0.05. Unless otherwise stated, we performed the paired T-tests in all comparisons. Linear regressions were plotted using the “ggpubr” package.

## Results

### Synthetic RNA from ATCC is a reliable positive control and reagent for standard curves

Having optimal positive controls in the form of standardized control RNA at a precisely defined concentration enables accurate quantification of viral loads from standard curves. While many vendors now provide synthetic SARS-CoV-2 RNA featuring gene targets recommended by the Centers for Disease Control and Prevention (CDC)^21^ and the German Centre for Infection Research (DZIF)^22^, preliminary studies have revealed that not all of them are at reliable concentrations^23^. Therefore, we tested two synthetic RNA preparations: one from the American Type Culture Collection (ATCC) and one from the United States National Institute of Standards and Technology (NIST) with listed concentrations of 10^5^ - 10^6^ copies/μl and 10^6^ copies/μl respectively. We chose these positive controls since they are easily accessible to other labs and are from reliable sources. A five-point ten-fold dilution series from a starting concentration of 10^4^ copies/μl to 10^0^ copies/μl was tested in duplicate ddPCR (Fig. 1a) and quadruplicate RT-qPCR (Fig. 1b) assays targeting the genes for the Envelope protein (E), Nucleocapsid proteins (N1, N2) and RNA dependent RNA Polymerase protein (RdRP)^21, 22^. Notably, the NIST standard was provided in two fragments, with fragment 1 bearing the E, N1 and N2 genes, and fragment 2 the RdRP gene^24^. The dilution series prepared by two different users working with independent aliquots of the standards revealed ATCC’s synthetic RNA standard to be a reliable control with high concordance across reactions. ddPCR, which allows for absolute quantification, revealed the starting concentration of these standards to be 10^6^ copies/μl. While the NIST standards also performed with high concordance within replicates, the concentration of fragment 2 was consistently found to be lower than the stated concentration by an order of magnitude. Further, one out of eight of the RT-qPCR reactions assaying the NIST RNA for the E gene at 10^4^ RNA concentration failed to amplify, likely due to an experimental error in the RT-qPCR assay. This result highlights the importance of running RT-qPCR assays in replicates. Given the reliable performance of the synthetic SARS-CoV-2 RNA from ATCC across both ddPCR and RT-qPCR assays testing 4 target genes, we decided to use this reagent across this study (Supplementary Fig. 1a).

**Fig. 1.**
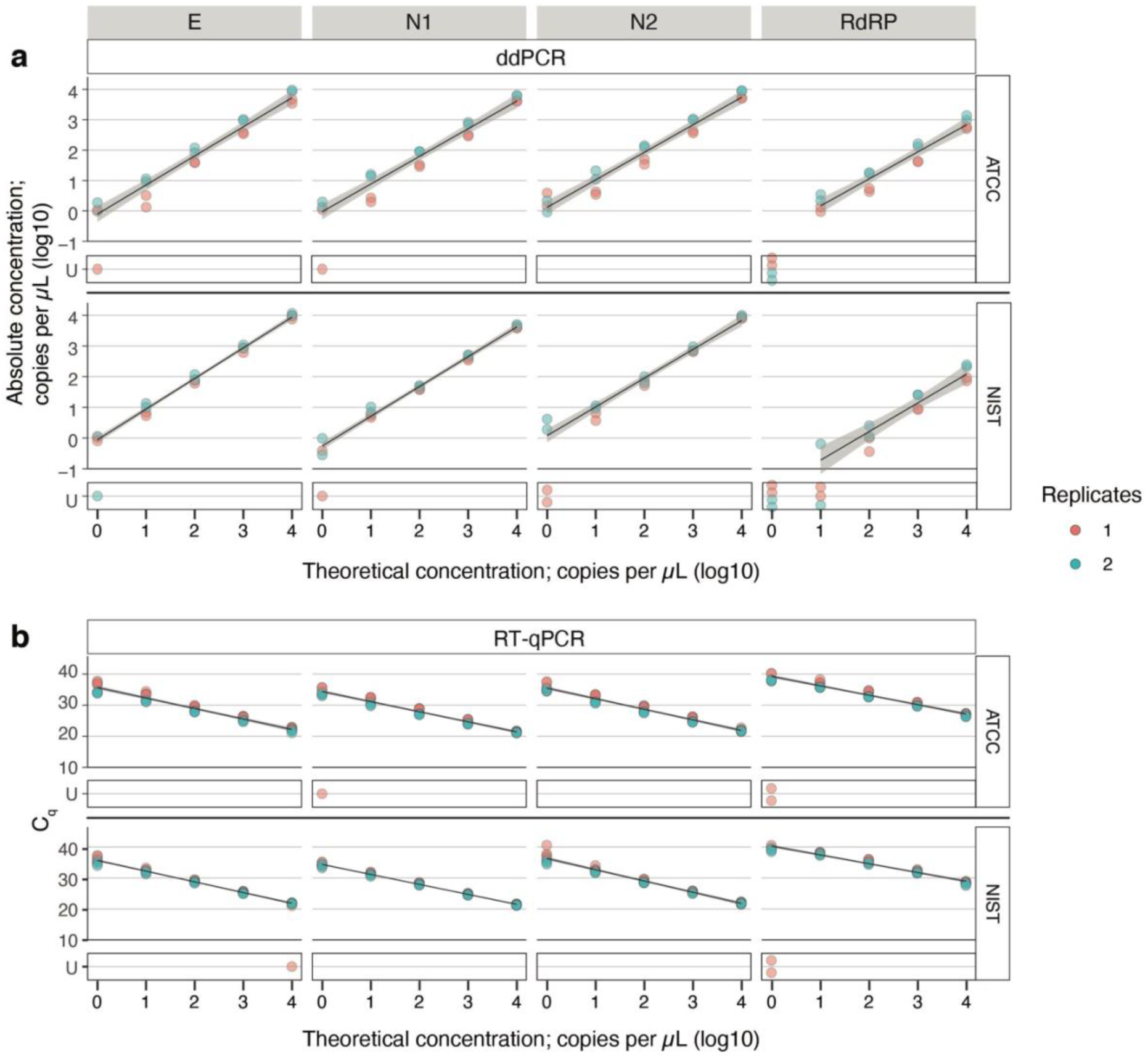
Robustness of synthetic SARS-CoV-2 RNA standards from ATCC and NIST. ddPCR and RT-qPCR assays targeting four SARS-CoV-2 RNA targets (E, N1, N2, and RdRP) across a five-point ten-fold concentration range of RNA standards from either ATCC or NIST (indicated on the tab to the right). **a**, The theoretical concentrations of RNA are plotted on the x-axis and absolute copy number derived from ddPCR is plotted on the y-axis. All assays were performed in duplicate. **b**, The theoretical concentrations of RNA are plotted on the x-axis and Cq derived from RT-qPCR is plotted on the y-axis. All assays were performed in quadruplicate. Replicates in red and blue refer to two independent experiments performed by two users using separate aliquots of samples. Linear regression is plotted in black and 95% confidence interval is shaded in gray. Samples that did not amplify are delineated as ‘U’ for undetermined and not included in the linear regression analysis. Associated statistics are summarized in Supplementary Table 1 and raw data is provided in Supplementary Information 1.

### ddPCR and RT-qPCR assays targeting the N1 gene are reliable means of estimating viral RNA concentration

We found all four primer/probe sets performed comparably in both the ddPCR and RT-qPCR assays based on accuracy of detection with respect to theoretical input concentration (Fig. 1a,b, Supplementary Table 1). Recent work has revealed N1 to be marginally more sensitive at detecting viral RNA from stool (manuscript in preparation). Therefore, we decided to target N1 in the rest of this study (Supplementary Fig. 1a). Given the high degree of concordance across replicate ddPCR and RT-qPCR reactions, we averaged results from replicate reactions in subsequent experiments. Further, since ddPCR allows absolute quantification of viral loads with high sensitivity^25^, while RT-qPCR is a more accessible platform for nucleic acid detection, we employed both techniques across the study to be more widely informative. In both assays, we used the one-step format that combines the reverse transcription and amplification steps in a single reaction for a simpler protocol.

### Standardized stool samples reveal that preservatives are important and that the ZV extraction kit performs best

We tested three different strategies to identify suitable methods of stool preservation for detection of SARS-CoV-2 viral RNA: a) stool stored without any preservative and resuspended in PBS (PBS), b) stool preserved in the OMNIgene-GUT tube (OG; DNA Genotek), a commonly used preservation kit in microbiome studies^26^, and c) stool preserved in the Zymo DNA/RNA shield collection kit (ZY; Zymo Research) that is explicitly rated for RNA preservation and virus inactivation.

In parallel, we also tested how these preservation methods interact with three different extraction kits - a) MagMAX viral/pathogen nucleic acid isolation kit (MM; Applied Biosystems), a magnetic-bead based protocol that has been successfully used with respiratory samples^27^, b) QiaAMP viral RNA minikit (QA; Qiagen), a column-based protocol that is used in many studies of fecal SARS-CoV-2 RNA^10–12^, c) Quick-RNA viral kit (ZV; Zymo Research), another column based protocol that is rated to be compatible with the ZY stool collection kit. All three of these extraction kits are scalable to a high-throughput format and therefore easily adaptable to clinical laboratories and other large-scale efforts.

In order to test and compare all combinations of preservation and extraction methods, we used standardized stool aliquots from NIST. Briefly, these are stool samples collected from a cohort of healthy, omnivorous human donors, which are then homogenized and made available in a ten-fold diluted format^16^. We spiked in synthetic SARS-CoV-2 RNA from ATCC (CoV-2 RNA) at two different concentrations (10^3^ and 10^4^ copies/μL of preserved stool sample) in this standardized stool sample and tested the combination of stool preservation and extraction kits to benchmark their performances across multiple target RNA concentrations (Supplementary Fig. 2a). Finally, RNA extractions were performed by two independent users, each in technical duplicates in order to guard against artefacts both across batches by the same user and across users.

Among the stool preservatives, more RNA was detected in ZY than OG in both samples spiked with 10^3^ and with 10^4^ concentrations of CoV-2 RNA when paired with the MM (Paired T-test; *P*^10^*^^3^* = 0.020, *P*^10^*^^4^* = 0.006) and QA (Paired T-test; *P*^10^*^^3^* = 0.006, *P*^10^*^^4^* = 0.049) extraction kits (Fig. 2a, Supplementary Table 2). Notably, ZV appears to efficiently isolate RNA across both preservatives. Next we compared the performance of the three extraction kits.

**Fig. 2.**
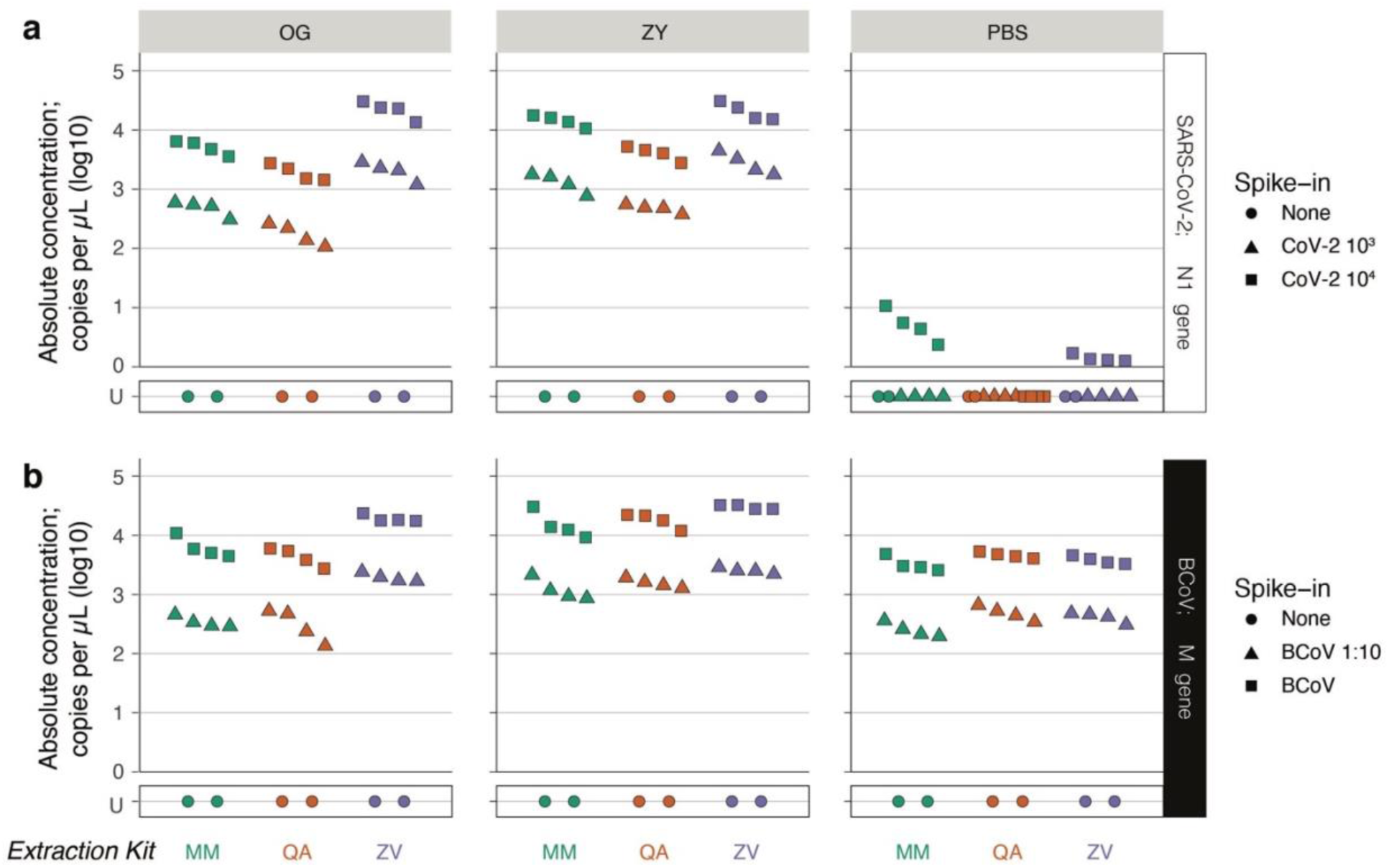
Efficacy of preservation and RNA extraction of SARS-CoV-2 and BCoV RNA from standardized NIST stool by ddPCR. Stool samples collected from omnivorous donors and processed into a single standardized matrix by NIST was spiked with ATCC CoV-2 RNA or BCoV vaccine. Spiked stool was preserved in the OMNIgene-GUT kit (OG), Zymo DNA/RNA shield buffer (ZY) and PBS (as indicated in the tab on the top). RNA was extracted from these samples by two independent users, each in duplicate, using the MagMAX Viral/Pathogen Kit (MM; green), QIAamp Viral RNA Mini Kit (QA; orange) or Zymo Quick-RNA Viral Kit (ZY; purple) as indicated on the x-axis. RNA was assayed using ddPCR. **a**, Absolute concentration of SARS-CoV-2 RNA assayed by ddPCR targeting the N1 gene is plotted on the y-axis. NIST stool matrix was spiked with 10^3^ (▲) or 10^4^ (∎) copies of ATCC synthetic SARS-CoV-2 RNA. **b**, Absolute concentration of BCoV RNA assayed by ddPCR targeting the M gene is plotted on the y-axis. NIST stool matrix was spiked with 1:10 diluted (▲) or undiluted (∎) BCoV vaccine. Control samples with no spiked in RNA (none; ●) were included in duplicate to estimate LoB. ‘U’ stands for undetermined and marks samples with no detectable RNA above LoB. Raw data provided in Supplementary Information 1.

Focusing our attention on the OG and ZY stool preservatives - in OG preserved samples, ZV outperforms MM by yielding more detectable RNA both in samples spiked with 10^3^ and with 10^4^ concentrations of CoV-2 RNA (Paired T-test; *P*^10^*^^3^* = 0.017, *P*^10^*^^4^* = 0.014), while in ZY preserved samples both kits perform comparably. Across both stool preservatives, MM and ZV outperform QA (Fig. 2a, Supplementary Table 2). Therefore, in the condition of standardized NIST stool samples spiked with two different concentrations of SARS-CoV-2 RNA we find that the ZY preservative and ZV extraction kit outperform the alternatives.

Notably, in the PBS preservative we detected SARS-CoV-2 RNA at roughly three orders of magnitude lower in eluates extracted from stool spiked with 10^4^ copies per µL of sample compared to OG or ZY. Across stool spiked with 10^3^ copies per µL of sample, we failed to detect any target RNA from PBS-preserved samples. We believe this is because the unpackaged SARS-CoV-2 RNA was degraded by RNAses known to be present in stool. While these data suggest that OG and ZY buffers are critical to preserving naked RNA in stool, testing preservatives in the context of unpackaged SARS-CoV-2 RNA may not be representative of clinical samples. This is because we do not yet know whether SARS-CoV-2 RNA shed in stool is in its naked unpackaged state, protected in a encapsulated structure (such as the virus itself, virus-like particles or host-double membrane vesicles), or a combination thereof.

Hence, we sought to identify a proxy for SARS-CoV-2 that is not known to cause disease in humans and is thus safe to handle in the laboratory at biosafety level 1. We picked Bovine coronavirus (BCoV), a virus that belongs to the same genus as SARS-CoV-2, *Betacoronovirus*, in the subgenus *Embecovirus*, sharing this taxonomy with other human pathogens (HCoV-HKU1 and HCoV-OC43). BCoV and SARS-CoV-2 share a common structural architecture, and are both positive stranded RNA viruses. Further, BCoV can be procured as an over-the-counter attenuated vaccine. Prior to stool based testing, we evaluated the performance of the ddPCR and RT-qPCR assays with the recommended primer/probe set to detect the M gene in BCoV RNA. Towards this, we used RNA extracted directly from the attenuated BCoV vaccine prepared in PBS in the absence of stool. We found both the ddPCR and RT-qPCR assays reliably tracked the seven-point ten-fold dilution of the RNA extracts (Supplementary Fig. 3a,b). Therefore, we next set out to test the same set of stool preservation and viral RNA extraction methods with the standardized NIST stool samples spiked with BCoV. To assess preservative and extraction kit performance across multiple target concentrations, we spiked BCoV both in its undiluted form and at a ten-fold dilution of the stock.

In this experiment, we recovered the target BCoV RNA even in PBS albeit at lower concentrations compared to other preservation methods (Fig. 2b). Further, the performance of the OG and ZY stool preservatives and the three RNA extraction kits were consistent with previous observations. Briefly, ZY preservative performs better than OG and PBS in all tested extraction kits, yielding more detectable target RNA (Fig. 2b). This observation is statistically significant (Supplementary Table 2) in all cases except in samples with the highest BCoV spike-in where MM performs comparably across the three preservatives. Given the superior performance of the ZY preservative, we went on to analyze how the three extraction methods fared in this condition. Here, ZV surpasses MM and QA at extracting BCoV RNA at both spike-in concentrations, though the difference between ZV and MM is not statistically significant in samples with the highest concentration of spiked in BCoV.

Alongside efforts to extract BCoV RNA from spiked stool samples, each user also extracted RNA directly from the BCoV vaccine without any stool sample. This allows us to evaluate if the extraction kits interact differently with encapsulated RNA, and also serves as a positive control for the extractions. Notably, we find that all extraction kits perform comparably, and reliably extract RNA from the BCoV vaccine (Supplementary Fig. 2b).

Finally, we sought to verify our observations using the more commonly used RT-qPCR assay as well. Notably, the RT-qPCR assays (Supplementary Fig. 2c,d) validate trends we observe in the ddPCR assays, but some of the differences in performance we note in the ddPCR assay are no longer significant (Supplementary Table 2). We attribute this to the lower sensitivity of RT-qPCR over ddPCR^25^. All experiments included stool samples with no spiked-in RNA to establish a reliable limit of blank (LoB). RNA extracted from stool samples spiked with BCoV had to be diluted ten-fold to arrive at a concentration range accurately quantifiable by ddPCR. Similarly, extracts from BCoV vaccine without stool had to be diluted 100-fold. Finally, given the concordance of results from biological replicates from the same user, we limited the number of replicates to one per user in subsequent experiments.

Taken together, in the NIST omnivore aqueous stool matrix, ZY best preserves both the SARS-CoV-2 naked RNA and encapsulated RNA from BCoV, a SARS-CoV-2 like *Betacoronovirus* (Supplementary Fig. 1b). Further, the ZV extraction kit is also the best performer across both these sample types. Finally, RNA, both in its unpackaged form and when packaged in a virus is susceptible to loss in PBS without any preservative.

### ZY preservative and QA extraction kits are broadly more effective in non-standardized stool samples

While the NIST stool samples were a useful, standardized preparation, this processed, pooled and diluted standardized stool sample is limited in its representation of regular clinical specimens. Therefore, we next tested the combinations of the preservatives and viral RNA extraction kits using undiluted and unprocessed stool samples from healthy donors, spiked with the SARS-CoV-2 RNA and BCoV standards. We picked the lower concentrations of both the SARS-CoV-2 RNA (10^3^) and BCoV (1:10 dilution) from our previous analysis to challenge the sensitivity of the combinations of preservation, extraction and detection techniques tested here.

We acquired stool samples from two healthy stool donors, one on an omnivorous diet (Omni) and the other on a vegetarian diet (Veg) (Supplementary Fig. 4a). Across conditions, the concentrations of target RNA detected from these matrices were lower than those from the NIST samples by around an order of magnitude (Fig. 2a, 3a). We used ddPCR to assay the performance of the preservatives and observed that in samples spiked with SARS-CoV-2 RNA, ZY yields more detectable RNA than OG when paired with both the MM (Paired T-test; *P* = .048) and QA (Paired T-test; *P* =.035) extraction kits (Fig. 3a, Supplementary Table 3). In the best performing preservative, ZY, all extraction kits perform comparably. Notably, PBS continues to perform poorly, yielding no detectable target RNA in all but one extraction. These results based on unprocessed non-standardized stool samples suggest that it is best to preserve samples in the ZY buffer, and that in this preservative, all three extraction kits can be used with comparable results.

**Fig. 3.**
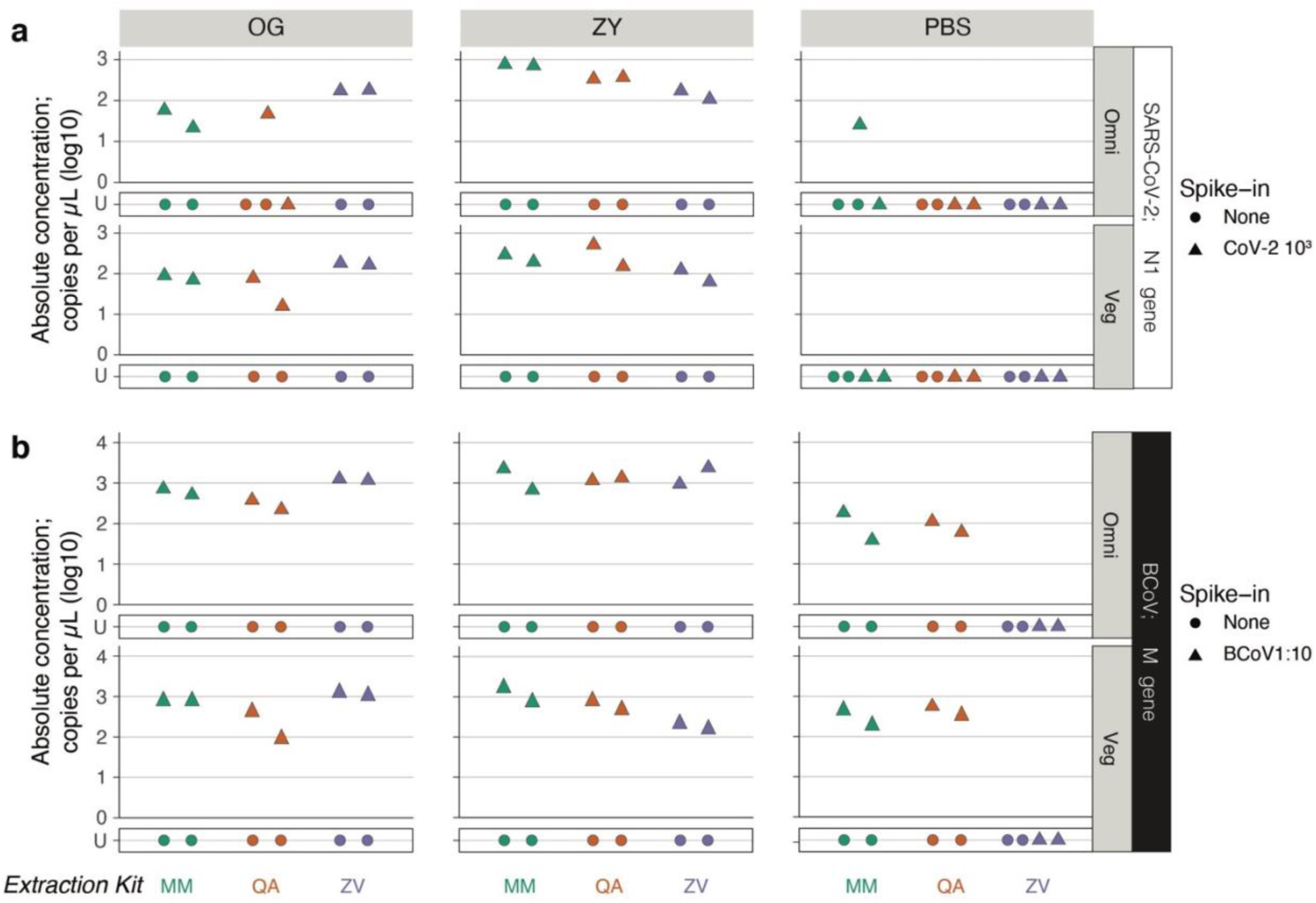
Evaluating preservation and extraction of SARS-CoV-2 and BCoV RNA from non-standardized stool samples using ddPCR. Stool samples were collected from healthy omnivorous (Omni) and vegetarian (Veg) donors and spiked with ATCC CoV-2 RNA or BCoV vaccine. Spiked stool was preserved in the OMNIgene-GUT kit (OG), Zymo DNA/RNA shield buffer (ZY) and PBS (as indicated in the tab on the top). RNA was extracted from these samples by two independent users using the MagMAX Viral/Pathogen Kit (MM; green), QIAamp Viral RNA Mini Kit (QA; orange) or Zymo Quick-RNA Viral Kit (ZY; purple) as indicated on the x-axis. RNA was assayed using ddPCR. **a**, Absolute concentration of SARS-CoV-2 RNA assayed by ddPCR targeting the N1 gene is plotted on the y-axis. Healthy stool samples were spiked with 10^3^ (▲) copies of ATCC synthetic SARS-CoV-2 RNA. **b**, Absolute concentration of BCoV RNA assayed by ddPCR targeting the M gene is plotted on the y-axis. Healthy stool samples were spiked with 1:10 diluted (▲) BCoV vaccine. Control samples with no spiked in RNA (none; ●) were included in duplicate to estimate LoB. ‘U’ stands for undetermined and marks samples with no detectable RNA above LoB. Raw data provided in Supplementary Information 1.

In the case of RNA encapsulated in BCoV, the two preservatives, OG and ZY, perform comparably in all but one instance; ZY offers an advantage in samples spiked with diluted BCoV (1:10) and extracted with QA (Paired T-test; *P* =.034) (Fig. 3b, Supplementary Table 3). Next, focusing on samples preserved in ZY, all three extraction kits yield comparable amounts of detectable viral RNA. Further, control extractions, with only the BCoV vaccine without any stool included in this batch also yielded comparable amounts of RNA across kits (Supplementary Fig. 4b). We note that RNA extracted from stool samples spiked with BCoV had to be diluted ten-fold to arrive at a concentration range accurately quantifiable by ddPCR, and those from BCoV vaccine without stool had to be diluted 100-fold.

Unlike the previous experiment with standardized diluted NIST stool, in this set of samples based on unprocessed healthy stool, we observe differences in the performance of ddPCR and RT-qPCR assays (Supplementary Fig. 4c,d). Interestingly, we detected BCoV from the PBS sample extracted with the ZV kit in the RT-qPCR assay, albeit at a high C_q_ value, but not in the ddPCR assay. This is one exception among all the assays performed in this study, and likely a false positive. Next, by and large, we were unable to detect the BCoV target in RNA extracted using the MM kit. This observation makes us suspect that PCR inhibitors are being coeluted with RNA when using the MM kit. Given these observations, we conclude that QA performed most reliably at yielding detectable RNA from BCoV spiked into non-standardized stool.

Overall, these experiments comparing the performance of preservatives and extraction kits on non-standardized stool samples revealed that ZY yields more detectable target RNA than OG and PBS (Supplementary Fig. 1b). Further, we are still unable to detect RNA from samples stored in PBS when trying to recover the unpackaged ATCC synthetic RNA spiked into stool.

Finally, while all extraction kits perform comparably at extracting unpackaged RNA, QA performs more reliably than MM and ZY at extracting BCoV encapsulated RNA.

### ZY collection and preservation is more effective than OG in the hands of patients

Experiments so far studied defined stool samples spiked in with a known amount of target RNA, and transferred to collection kits in a precise, controlled environment. This is useful towards testing kits head-to-head. However, in reality, stool samples are likely to be collected by patients or health-care practitioners outside of well-controlled laboratory spaces. Therefore, it is important to compare the performance of the OG and ZY stool preservatives in this practical use-case setting.

To this end, we leveraged a recent large-scale study that captured the dynamics of fecal SARS-CoV-2 viral RNA shedding (unpublished data, manuscript in preparation)^17^. Briefly, this study collected stool samples from COVID-19 outpatients that were enrolled in a clinical trial of Peginterferon Lambda-1a in both the OG and ZY preservatives. Further, they extracted RNA using QA and assayed viral load using RT-qPCR.

From this data set, we picked instances of paired OG and ZY viral loads determined from samples collected from the same patient at the same time. Out of 172 such samples, 82 did not yield a detectable amount of target RNA in either preservative and were left out of further analysis. Taking the 90 paired samples in which we detected the viral RNA targets in at least one of the preservatives, we plotted their log_10_-transformed concentrations in a scatter plot (Fig. 4). Here we fitted a linear regression, excluding samples that yielded RNA in only one of the two preservatives since these skewed the regression. Notably, 29 of these paired samples yielded detectable RNA only in ZY, in comparison to 7 in only OG. The linear regression from the paired samples stored in OG and ZY reveals that among samples for which both samples tested positive, OG samples had a roughly 60% lower detected concentration of RNA. Finally, we also calculated the mean of the differences between the log_10_-transformed viral RNA concentrations from these paired samples, including ones that were only detected in one of the two preservatives. This revealed that ZY preserved samples yielded more RNA by 0.477 log_10_ units (or ∼3 times more) (Independent T-test; *P =* 2.36E-07).

**Fig 4.**
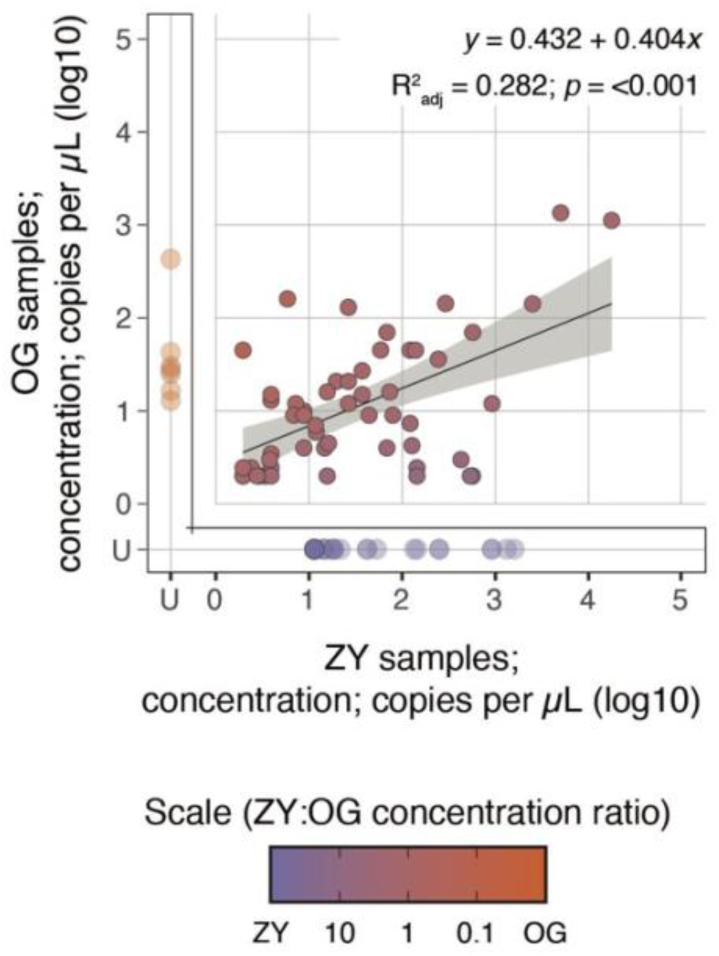
Relationship between yields of SARS-CoV-2 RNA extracted from clinical samples stored in two different preservatives. Paired stool samples were collected in the OMNIgene-GUT kit (OG) and Zymo DNA/RNA shield buffer (ZY) preservatives from COVID-19 outpatients enrolled in a clinical trial of Peginterferon Lambda-1a. RNA from these samples had been extracted using the QIAamp Viral RNA Mini Kit (QA), assayed by RT-qPCR targeting the N1 gene and previously reported. Scatter plot of the reported concentrations of paired stool samples with concentrations derived from the ZY preserved samples on the x-axis and from the OG preserved samples on the y-axis. Linear regression is plotted in black and 95% confidence interval is shaded in gray. Samples that are reported to not have amplified are delineated as ‘U’ for undetermined and not included in the linear regression analysis. This breaks down as 29 samples that were detected only in the ZY preservative, 7 that were detected only in the OG preservative and 54 that were detected in both. Associated regression equation and statistics are inset on the top right. Color gradient from purple to orange represents the ratio of the concentration of RNA derived from the ZY preserved sample (purple) to that derived from the OG preserved sample (orange). Raw data are provided in Supplementary Information 1.

While the ZY preservative may be more effective at protecting RNA, it is also possible that the ZY collection kit ends up with more stool compared to the OG kit. In order to address this question we estimated how much of the sample from either of these kits is actually composed of stool. To this end, we randomly selected paired samples collected in the OG and ZY tubes from the biobank of stool samples collected from COVID-19 outpatients enrolled in the aforementioned clinical trial of Peginterferon Lambda-1a (manuscript in preparation).

Specifically, each of these pairs was collected from the same patient at the time of enrollment in the study. We took two biopsy punches from each of these 10 stool samples and measured their wet weight. Next, we dried these samples on a heat block for 72 hours and measured their dry weight. The percentage of dry weight to wet weight represents the proportion of patient stool biomass in the original sample. We found that 31.4 ± 1.6% of sample weight in the ZY preservative corresponds to stool biomass, compared to 13.6 ± 3.3% of sample weight in the OG preservative (Table 2; Paired T-test; *P* = 5.49E-6). Remarkably, this roughly 3-fold difference in stool biomass tracks closely with the 3-fold difference we observe above in the performance of the two kits. Therefore, likely, the two kits preserve and yield comparable amounts of detectable SARS-CoV-2 RNA, when accounting for the amount of input stool.

**Table 2.** Measurement of wet weight and dry weight from 5 paired stool samples collected from COVID-19 patients in OG and ZY.

| **Sample ID** | **ZY** |  |  | **OG** |  |  |
| --- | --- | --- | --- | --- | --- | --- |
|  | **Wet weight** | **Dry weight** | **Percent stool biomass (%)** | **Wet weight** | **Dry weight** | **Percent stool biomass (%)** |
| Lambda_248 | 0.119 | 0.036 | 30.252 | 0.172 | 0.031 | 18.023 |
| Lambda_327 | 0.145 | 0.043 | 29.655 | 0.117 | 0.011 | 9.402 |
| Lambda_223 | 0.101 | 0.034 | 33.663 | 0.168 | 0.023 | 13.690 |
| Lambda_264 | 0.096 | 0.031 | 32.292 | 0.117 | 0.018 | 15.385 |
| Lambda_292 | 0.157 | 0.049 | 31.210 | 0.111 | 0.013 | 11.712 |
| **Average** |  |  | 31.414 |  |  | 13.642 |
| **Std deviation** |  |  | 1.606 |  |  | 3.314 |

However, the difference in stool biomass across the two kits is surprising to us, since reading the manufacturer’s instructions suggests that the OG kit would end up with a marginally higher concentration of stool. In fact, the experiments with stool from NIST and healthy donors described in this work followed these instructions and added 500 mg of stool to OG (containing 2 ml of buffer), and 1000 mg of stool to ZY (containing 9 ml of buffer). We suspect that this difference in stool input we observe in the clinical samples may be the effect of the format of the two kits. Specifically, the OG kit is composed of a specific receptacle of defined volume to collect stool, while the ZY kit is just a standard collection tube with a proprietary buffer (Supplementary Fig. 5). The ZY kit has plenty of room in the tube above the buffer level, so study subjects may have been inclined to load more stool in the ZY kit.

Taken together, we find that the ZY kit yields more detectable RNA than the OG kit both with samples prepared in strictly controlled experimental conditions carried out in the laboratory, and on those collected in the field by patients (Supplementary Fig. 1c). This superior performance may be the result of a better preservative, differential usage of these kits, or a combination thereof.

### Comparing the performance of extraction kits on clinical samples collected in the ZY preservative

Given the superior performance of the ZY preservative in both standardized and clinical samples, we next tested how the three extraction kits perform with “real life” clinical samples preserved in this modality.

In order to test the extraction kits, we picked 5 random samples from the Peginterferon Lambda-1a (unpublished data, manuscript in preparation) biobank that were collected in ZY on the day of enrollment, when we expect a higher fecal viral load. Recognizing that there is a high cost to false negatives in the detection of SARS-CoV-2 viral RNA in stool samples, we incorporated a reliable control to track efficiency of RNA extraction, without compromising the yield of the target SARS-CoV-2 RNA. Hence, we took two aliquots of the stool from each of the chosen 5 clinical samples, and spiked undiluted BCoV in one of them as a reference extraction control (Supplementary Fig. 6a). We then extracted RNA from the 10 stool samples using the three extraction kits.

Looking at the stool samples with spiked BCoV, we find that QA may perform marginally better than MM and ZV yielding more detectable target RNA, albeit not to statistical significance (Supplementary Fig. 1c, Supplementary Table 4). Comparing the yield of SARS-CoV-2 RNA in samples with (Fig. 5a) and without (Supplementary Fig. 6b) spiked in BCoV reveals that the addition of this control does not significantly affect the yield of RNA (Fig. 5b) with the exception of sample # 4 extracted using QA. Here, inexplicably, we find that the stool sample with spiked BCoV yields SARS-CoV-2 RNA while the unspiked sample does not. We suspect this to be an anomaly warranting further exploration with a larger sample set. However, it appears reasonable to conclude that addition of BCoV does not negatively impact the extraction and detection of SARS-CoV-2 RNA. We also report the detected copies of N1 per gram of stool to normalize the copies of SARS-CoV-2 RNA to the amount of stool placed into the preservative by patients. This does not alter our conclusions regarding the best extraction kit, but given the differing input of stool to various preservative options, we believe that reporting detected copies per gram of stool where possible will best harmonize reported viral loads of SARS-CoV-2 in feces (Supplementary Information 2).

**Fig. 5.**
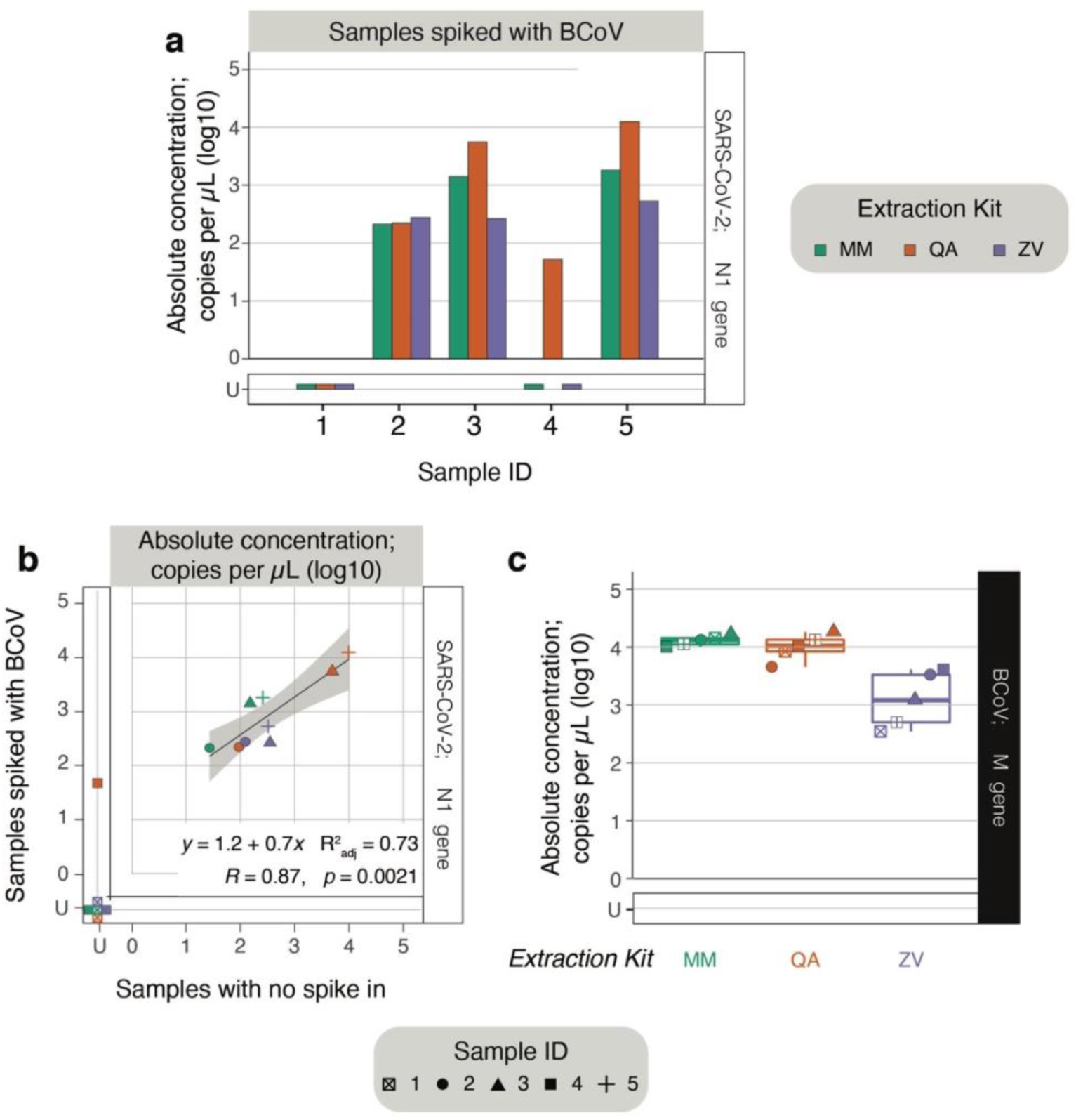
Testing efficiency of three extraction kits using clinical samples stored in the ZY preservative and spiked with BCoV. Stool samples were collected in the Zymo DNA/RNA shield buffer (ZY) preservative from five COVID-19 outpatients enrolled in a clinical trial of Peginterferon Lambda-1a. All samples were spiked with 10 μl of undiluted BCoV vaccine. In parallel, the same set of samples were processed without any spike-in. RNA from these samples were extracted using the MagMAX Viral/Pathogen Kit (MM; green), QIAamp Viral RNA Mini Kit (QA; orange) or Zymo Quick-RNA Viral Kit (ZY; purple). **a.** RNA from samples with BCoV spiked in were assayed for SARS-CoV-2 RNA using ddPCR targeting the N1 gene. Anonymized sample identities are listed on the x-axis and absolute concentration is listed on the y-axis. **b.** Scatter plot of the absolute concentration of SARS-CoV-2 RNA derived from samples without any spike-in (x-axis) versus those with 10 μl of undiluted BCoV spiked in (y-axis), measured using ddPCR targeting the N1 gene. Linear regression is plotted in black and 95% confidence interval is shaded in gray. ‘U’ stands for undetermined and indicates samples without no detectable RNA above the LoB; these undetermined concentrations samples are not included in the linear regression analysis. Associated regression equation and statistics are inset on the bottom right. **c.** RNA extracted from samples with BCoV spiked were assayed for BCoV RNA using ddPCR targeting the M gene. Cumulative box plot of the absolute concentrations of BCoV RNA across the clinical samples marking the median, first and third quartile and 95% confidence interval. ‘U’ stands for undetermined and marks samples with no detectable RNA above LoB. Raw data provided in Supplementary Information 1.

We next analyzed the cumulative yield of BCoV RNA from each of these clinical samples and found that samples extracted with the MM and QA kits performed comparably and reliably, whereas BCoV RNA detection was more variable in samples extracted with the ZV kit (Fig. 5c). Therefore, we demonstrate here how an easily accessible, over-the-counter attenuated BCoV vaccine can be leveraged as a reliable spike in control.

All results considered, we recommend using the ZY preservative to collect stool samples, and the QA extraction method to purify SARS-CoV-2 RNA (Supplementary Fig. 7). In instances where variability in extracted RNA yield or coelution of polymerase inhibitors are anticipated, we suggest spiking in 10 μl of BCoV vaccine to 500 μls of stool prior to storage and extraction in order to guard against false negatives. We have validated here that BCoV serves as a reliable control and does not affect the yield of SARS-CoV-2 RNA.

## Discussion

Fecal shedding of SARS-CoV-2 RNA is emerging as a key manifestation of COVID-19 infection with vast implications for patient health and in the epidemiology of the disease.

However, methods to collect and preserve patient samples, and to extract viral RNA for the robust detection and quantification of SARS-CoV-2 remain underexplored. Therefore, we compare strategies for each of these steps in the testing of fecal samples and report here an optimized methodology. We have focused our efforts on reagents that are easily available and kits that are scalable to a high-throughput format, therefore enabling straightforward adoption for work in research and clinical laboratories.

We tested three different strategies for sample collection and preservation. First, the most common strategy involves collecting stool without any preservative^28–36^. These samples are resuspended in PBS for viral RNA extraction^10^. Next, we also tested the OMNIgene-GUT tube (OG) that is widely used in stool collection for gut microbiome analysis. Finally, we included the Zymo DNA/RNA Shield kit (ZY) as a sample preservation method that is explicitly marketed for RNA preservation. Across three different types of stool samples, ZY consistently performed better than OG and PBS, enabling both the recovery of naked, unpackaged SARS-CoV-2 RNA and BCoV RNA encapsulated in a *Betacoronavirus* similar to SARS-CoV-2. Most importantly, analysis of data from a large study of outpatients with mild to moderate COVID-19 further validated the conclusion that ZY was the most effective preservation method. Conclusions from our study in combination with existing evidence that SARS-CoV-2 RNA is susceptible to degradation from freezing stool samples without any preservative^11^ highlights the critical importance of storing stool samples in an appropriate buffer.

Next, we compared three different extraction kits for their potential to effectively isolate viral RNA. Two of these kits, the QIAamp Viral RNA Mini Kit (QA) and Zymo Quick-RNA Viral Kit (ZY) are column based kits, while the MagMAX Viral/Pathogen Kit (MM) is based on magnetic beads. We tested these kits by performing replicate nucleic acid extractions of stool samples prepared in the laboratory spiked with SARS-CoV-2 synthetic RNA or BCoV vaccine. Here we found that the performance of the extraction kits was influenced by the preservative, nature of stool and the target RNA. We focus our discussion on the performance of the extraction kits in combination with the best performing preservative, ZY. Here, we observe that ZV most effectively extracted both the unpackaged SARS-CoV-2 RNA and the packaged BCoV RNA from the standardized, diluted NIST stool samples. However, from non-standardized healthy stool samples and clinical samples, QA performed more consistently, yielding detectable viral RNA across conditions. Notably, while MM performed well in many of the experiments, we find preliminary evidence that this protocol may allow the co-purification of PCR inhibitors. We glean this observation from experiments performed with BCoV spiked into non-standardized healthy stool samples. Taken together, we recommend using the QA extraction kit in tandem with the ZY preservative as a strategy for the robust and sensitive detection of SARS-CoV-2 RNA from stool (Supplementary Fig. 7).

Stool based testing of SARS-CoV-2 offers unique applications in healthcare. With emerging evidence that prolonged shedding of SARS-CoV-2 RNA in stool may be linked to an improved immune response^7^, there may be an opportunity to leverage fecal testing of RNA as a prognostic marker. Further, if the limited evidence of possible oral-fecal transmission of SARS-CoV-2 proves true, our ability to reliably test stool samples would be vital to controlling the spread of the pandemic as well as to inform healthcare practices such as fecal microbiota transplants. Finally, this option protects healthcare practitioners from having to be in close proximity to patients during sample collection. In all of these applications, it is critical to incorporate strategies to mitigate false negatives. Such false negatives may arise from errors in sample preservation, RNA extraction and presence of inhibitors that affect detection through PCR-based methods. Therefore, in this study, we also evaluate potential controls to guard against instances of such false negatives. We find that the widely accessible, safe, BCoV vaccine can be effectively spiked into stool samples prior to storage and extraction. Recovery of BCoV RNA assayed by targeting the M gene serves as a reliable metric of variation across batches of RNA preparations without affecting the yield of SARS-CoV-2 RNA in the samples. We believe BCoV to be a valuable proxy for SARS-CoV-2 since they belong to the same genus, *Betacoronavirus*, and predominantly share viral architecture. Therefore, using BCoV as a spiked-in control will help gain confidence in negatives as true negatives rather than a result of experimental artifacts (Supplementary Fig. 7).

Further, given the clinical implications, it is equally important to avoid false positives in the detection of SARS-CoV-2 RNA in stool. To this end, it is vital to establish a limit of blank (LoB) with every batch of experiments. This allows the confident identification of true positive samples over experimental noise. Guidelines from the Clinical & Laboratory Standards Institute (CLSI; EP17-A) provide a roadmap for a rigorous evaluation of LoB^37^. We recognize that this is a high bar for non-clinical research laboratories to meet. Alternatively, as demonstrated here, including comparable control stool samples from NIST or healthy donors in every batch of viral RNA extraction and detection will also serve to boost confidence in the detection of SARS-CoV- 2 RNA as being a true positive (Supplementary Fig. 7).

Finally, it is important to be able to quantify the viral RNA load in stool as a potential indicator of the state and prognosis of infection in patients. To this end, while ddPCR provides a powerful platform capable of determining the absolute concentration of RNA, we recognize this may be cost prohibitive and inaccessible. Therefore, additionally, we demonstrate here experimental strategies that enable the adoption of the more accessible RT-qPCR assay to enable the accurate detection and relative quantification of viral load in samples (Supplementary Fig. 7). Lastly, given differing amounts of stool collected by every patient and in different experiments, we recommend reporting quantified viral RNA load in terms of copies per gram of stool. This enables a normalized dataset that will allow us to harmonize reported fecal viral loads of SARS-CoV-2 RNA across studies.

SARS-CoV-2 has been a deadly pathogen causing extensive morbidity and mortality. Given the current understanding of coronaviruses, it is likely that SARS-CoV-2 will not be the last virus of this nature to cause an epidemic. Further, many coronaviruses are capable of infecting the gastrointestinal tract. In this context, we hope that the current work helps create a roadmap for fecal testing of coronavirus infections enabling the robust detection and quantification of viral RNA in stool.

## Data Availability

All data included in this manuscript are available as supplementary data tables within this submission.

## Acknowledgements

We thank Jason Andrews, Nasa Sinnott-Armstrong and Renu Verma for guidance on processing stool samples and detection of RNA using RT-qPCR, Angela Rogers and Julie Parsonnet for providing stool and rectal swab samples from patients admitted at Stanford Hospital, Scott Jackson and Stephanie Servetas from NIST for providing us aliquots of standardized stool, the Applied Genetics Group at NIST for aliquots of the SARS-CoV-2 synthetic RNA, Dean Felsher for access to the QuantStudio 12K Flex qPCR machine, Yvonne Maldonado and Jonathan Altamirano for helping acquire funding to support towards this work, Said Attiya and Dhananjay Wagh for guidance on applying ddPCR assays, David Solor-Cordero for assistance setting up the Biomek FX and providing access, Luisa Jiminez and Sopheak Sim for assistance in using the Stanford Functional Genomics Facility and High-Throughput Bioscience Center. We are grateful to the Peginterferon-λ1a clinical trial team for coordinating procurement of stool samples from outpatients enrolled in this trial. Biorender has been a valuable resource to creating schematic illustrations. This work was supported by a ChemH-IMA grant and the Stanford Dean’s Postdoctoral Fellowship (A.N.). The Bhatt lab is supported by NIH R01 AI148623 and R01 AI143757. A.H. is supported by an NSF Graduate research fellowship program grant. A.N., A.H., S.Z., E.F.B., S.E.V., M.W., A.Bo., and A.S.B. are co-inventors on a U.S. provisional patent application #63/172,045 that has been filed and relates to the methods presented in this manuscript.

## Supplementary Figures

**Supplementary Fig. 1.**
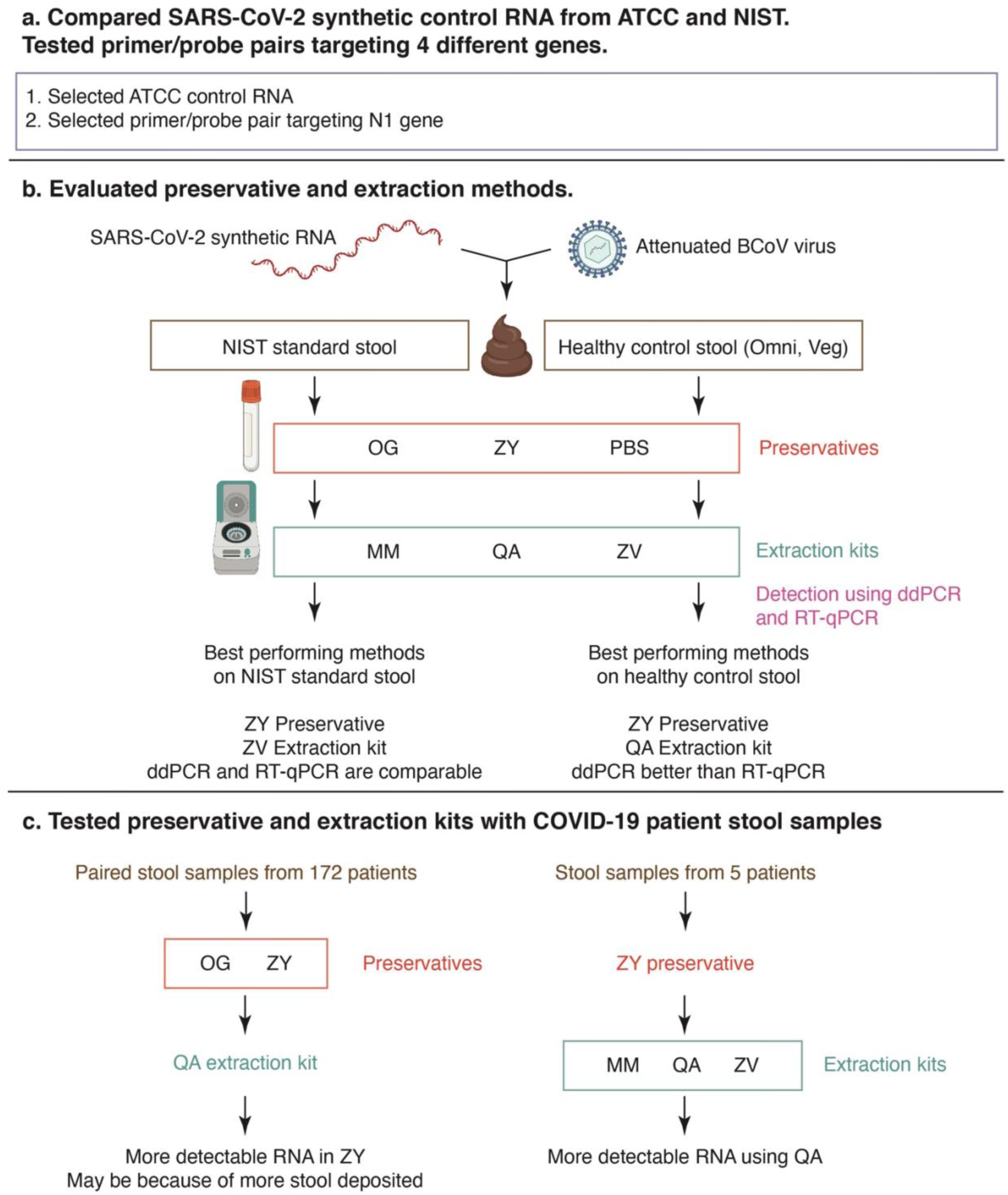
Schematic illustration of key experiments and results. **a.** Comparisons of SARS-CoV-2 synthetic control RNA standards from ATCC and NIST revealed that ATCC performed reliably. Further, the primer/probe set targeting the N1 gene in SARS-CoV-2 RNA performed well at detecting viral RNA. **b.** Standardized stool samples from NIST, and non-standardized stool samples from two healthy donors each on an omnivorous and vegetarian diet were employed to evaluate various preservatives and extraction methods. Stool samples were spiked with synthetic SARS-CoV-2 RNA or attenuated BCoV vaccine at defined concentrations. Samples were preserved in the OG, ZY or PBS buffers, and viral RNA was extracted using the MM, QA or ZY kits. Viral RNA was quantified using ddPCR and RT-qPCR targeting the N1 gene in SARS-CoV-2 or the M gene in BCoV. In NIST samples, ZY and ZV performed best while in healthy stool samples, ZY and QA performed best. Although both ddPCR and RT-qPCR performed comparably at detection RNA extracted from NIST samples, ddPCR performed better in RNA extracted from healthy stool samples. **c.** Performance of the preservatives and extraction kits were assessed using clinical samples. In the first experiment (left) viral RNA extracted using QA from 172 paired patient stool samples preserved in the OG and ZY buffers was analyzed. ZY preserved samples yielded more detectable RNA, perhaps because more stool was deposited in this kit. Next, in the second experiment (right) viral RNA was extracted from 5 patient stool samples preserved in ZY using MM, QA and ZV. Here, QA yielded more detectable RNA.

**Supplementary Fig. 2.**
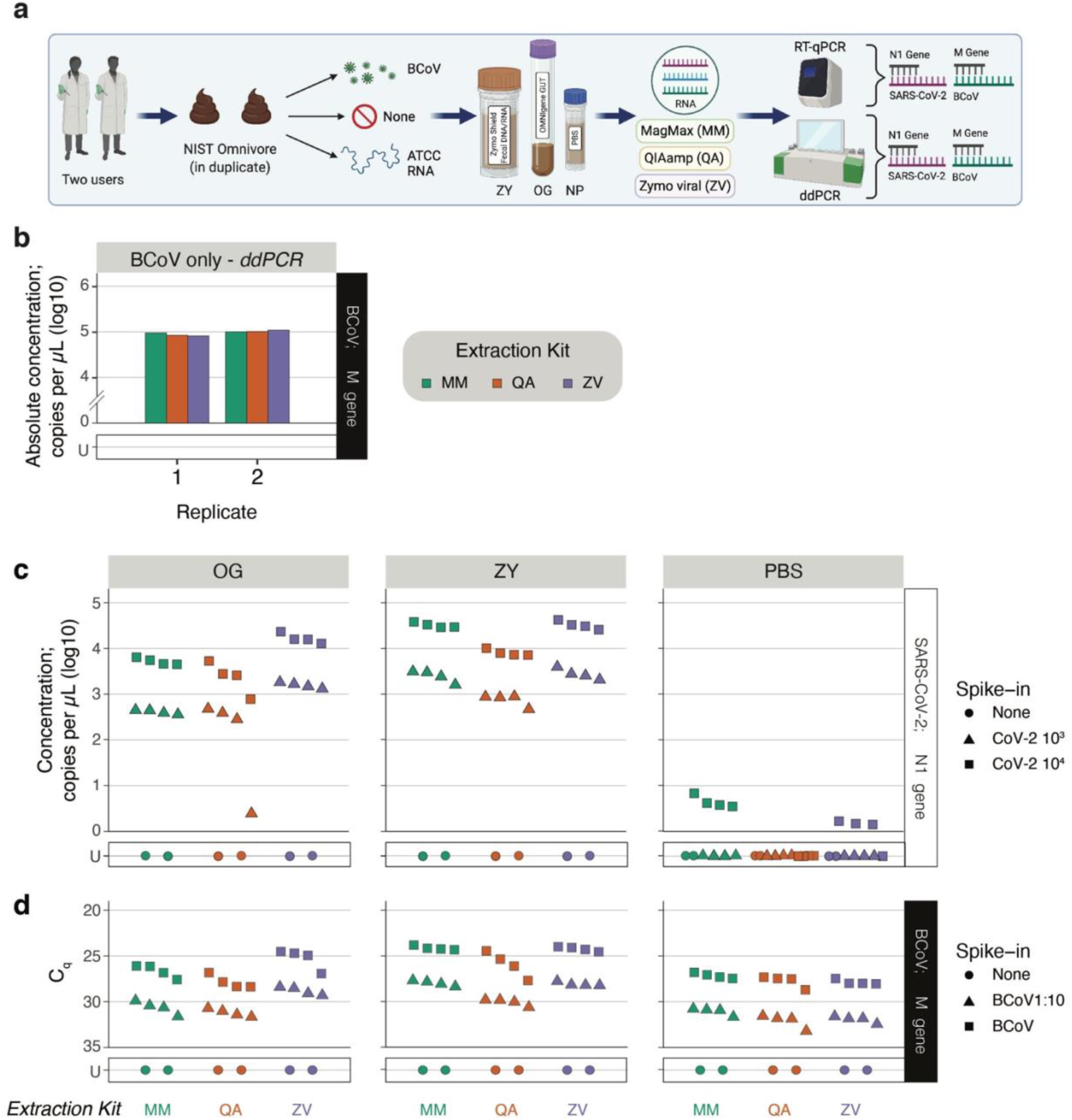
Efficacy of preservation and RNA extraction of SARS-CoV-2 and BCoV RNA from standardized NIST stool. **a.** Two independent users performed this experiment in duplicate. Stool samples collected from omnivorous donors and processed into a standardized matrix by NIST was spiked with ATCC CoV-2 RNA, a BCoV vaccine or equal volume of PBS (no RNA). Spiked stool was preserved in the OMNIgene-GUT kit (OG), Zymo DNA/RNA shield buffer (ZY) and PBS. RNA was extracted from these samples using the MagMAX Viral/Pathogen Kit (MM), QIAamp Viral RNA Mini Kit (QA) or Zymo Quick-RNA Viral Kit (ZY). RNA was assayed using ddPCR and RT-qPCR targeting the SARS-CoV-2 N1 gene or BCoV M gene. **b.** As an extraction control, RNA was isolated from the BCoV vaccine directly without the stool matrix using MM (green), QA (orange) and ZY (purple) kits. Each user included a set of these extractions (indicated in the x-axis). Absolute concentration of BCoV RNA assayed by ddPCR targeting the M gene is plotted on the y-axis. **c**, RNA extracted using the MM (green), QA (orange) and ZY (purple) kits are listed on the x-axis, and concentration of SARS-CoV-2 RNA assayed by RT-qPCR targeting the N1 gene is plotted on the y-axis. NIST stool matrix was spiked with 10^3^ (▴) or 10^4^ (◾) copies of ATCC synthetic SARS-CoV-2 RNA and processed in quadruplicate. **d**, RNA extracted using the MM (green), QA (orange) and ZY (purple) kits are listed on the x-axis, and Cq value of RT-qPCR assays targeting the BCoV M gene is plotted on the y-axis. NIST stool matrix was spiked with 1:10 diluted (▴) or undiluted (◾) BCoV vaccine. Control samples with no spiked in RNA (none; ⬤) were included in duplicate to estimate LoB. ‘U’ stands for undetermined and marks samples with no detectable RNA above LoB. RT-qPCR assays were run in technical duplicates and the mean values are represented here. Raw data provided in Supplementary Information 1.

**Supplementary Fig. 3.**
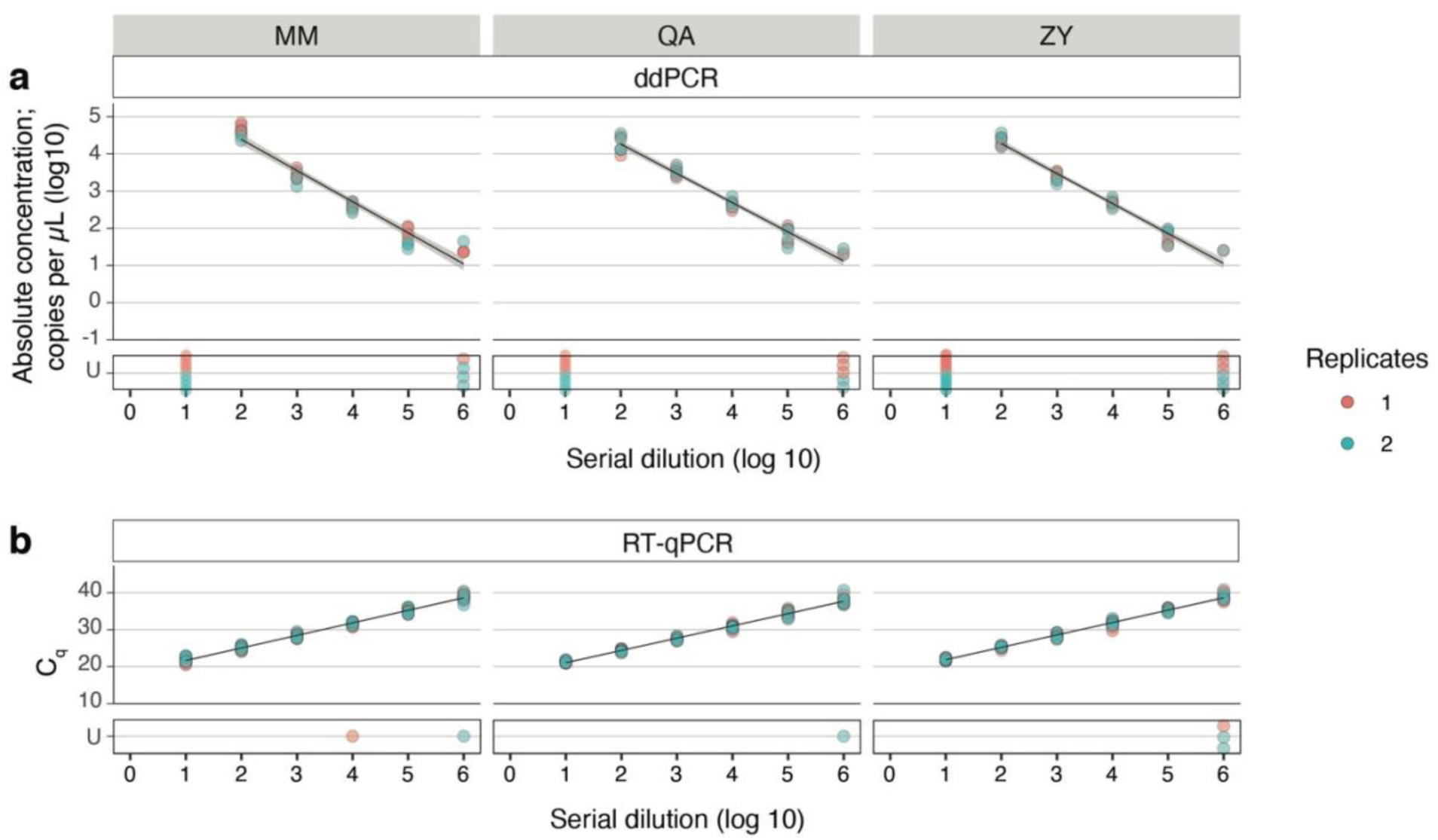
Robustness of primer/probe set at quantifying BCoV. ddPCR and RT-qPCR assays targeting M gene from BCoV across a seven-point ten-fold dilution series of RNA extracted from BCoV vaccine. RNA was extracted using either the MagMAX Viral/Pathogen Kit (MM), QIAamp Viral RNA Mini Kit (QA) or Zymo Quick-RNA Viral Kit (ZY) as indicated on the tab to the top. **a,** Dilutions of RNA are plotted on the x-axis and absolute copy number derived from ddPCR is plotted on the y-axis. All assays were performed in duplicate. **b**, Dilutions of RNA are plotted on the x-axis and Cq derived from RT-qPCR is plotted on the y-axis. All assays were performed in quadruplicate. Replicates in red and blue refer to two independent experiments performed by two users using separate extractions of RNA. Linear regression is plotted in black and 95% confidence interval is shaded in gray. Samples that did not amplify are delineated as ‘U’ for undetermined and not included in the linear regression analysis. Associated statistics are summarized in Supplementary Table 1 and raw data is provided in Supplementary Information 1.

**Supplementary Fig. 4.**
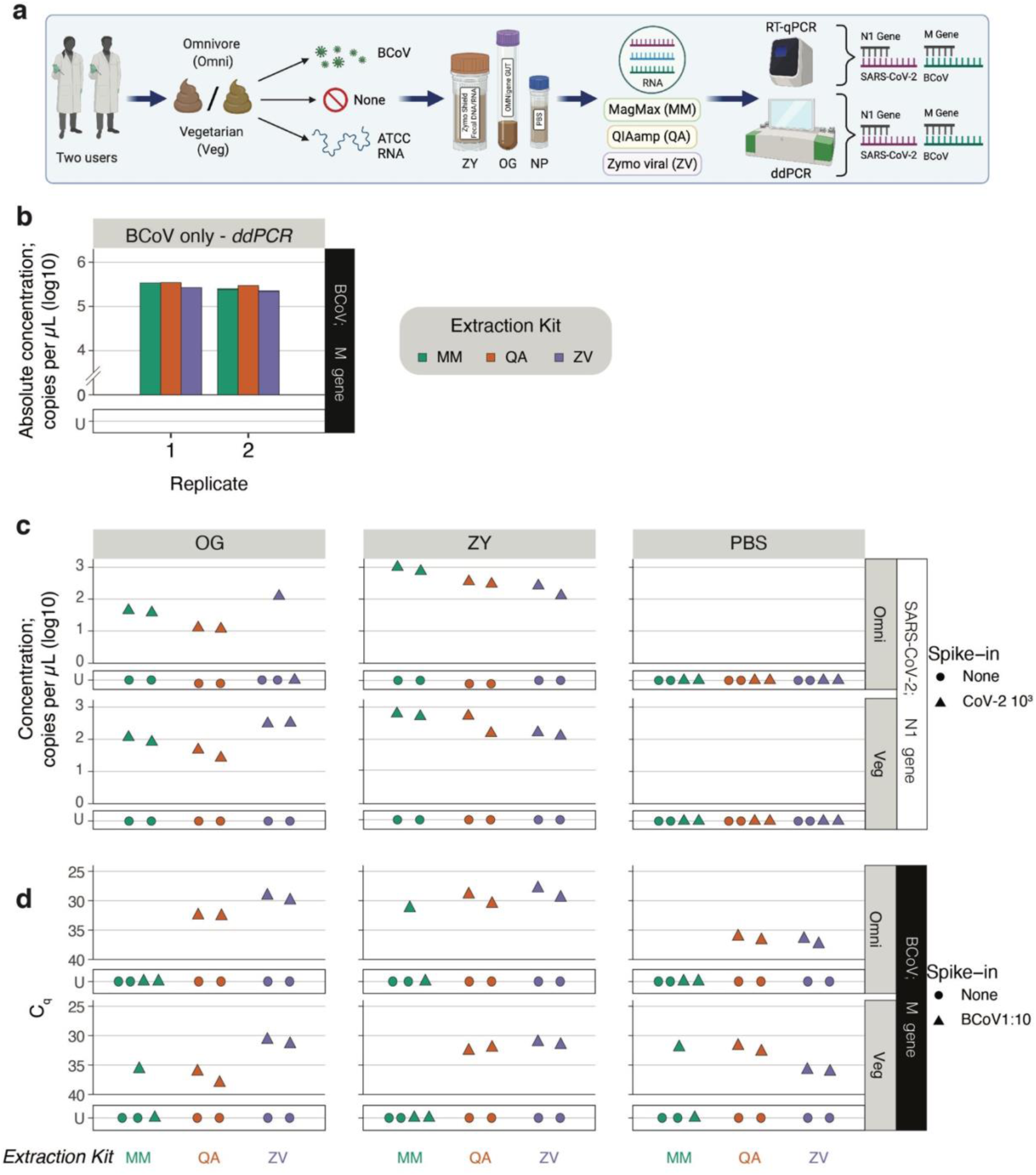
Performance of preservation and extraction of SARS-CoV-2 and BCoV RNA from non-standardized stool samples from healthy donors. **a.** Two independent users performed this experiment. Stool samples were collected from healthy omnivorous (Omni) and vegetarian (Veg) donors and spiked with ATCC CoV- 2 RNA or BCoV vaccine or equal volume of PBS (no RNA). Spiked stool was preserved in the OMNIgene-GUT kit (OG), Zymo DNA/RNA shield buffer (ZY) and PBS. RNA was extracted from these samples using the MagMAX Viral/Pathogen Kit (MM), QIAamp Viral RNA Mini Kit (QA) or Zymo Quick-RNA Viral Kit (ZY). RNA was assayed using ddPCR and RT-qPCR targeting the SARS-CoV-2 N1 gene or BCoV M gene. **b.** As an extraction control, RNA was isolated from the BCoV vaccine directly without the stool matrix using MM (green), QA (orange) and ZY (purple) kits. Each user included a set of these extractions (indicated in the x-axis). Absolute concentration of BCoV RNA assayed by ddPCR targeting the M gene is plotted on the y-axis. **c**, Concentration of SARS-CoV-2 RNA assayed by RT-qPCR targeting the N1 gene is plotted on the y-axis. Healthy stool samples were spiked with 10^3^ (▴) copies of ATCC synthetic SARS-CoV-2 RNA. **d**, Cq values from RT-qPCR assays of BCoV RNA targeting the M gene are plotted on the y-axis. Healthy stool samples were spiked with 1:10 diluted (▴) BCoV vaccine. Control samples with no spiked in RNA (none; ⬤) were included in duplicate to estimate LoB. ‘U’ stands for undetermined and marks samples with no detectable RNA above LoB. Raw data provided in Supplementary Information 1.

**Supplementary Fig. 5.**
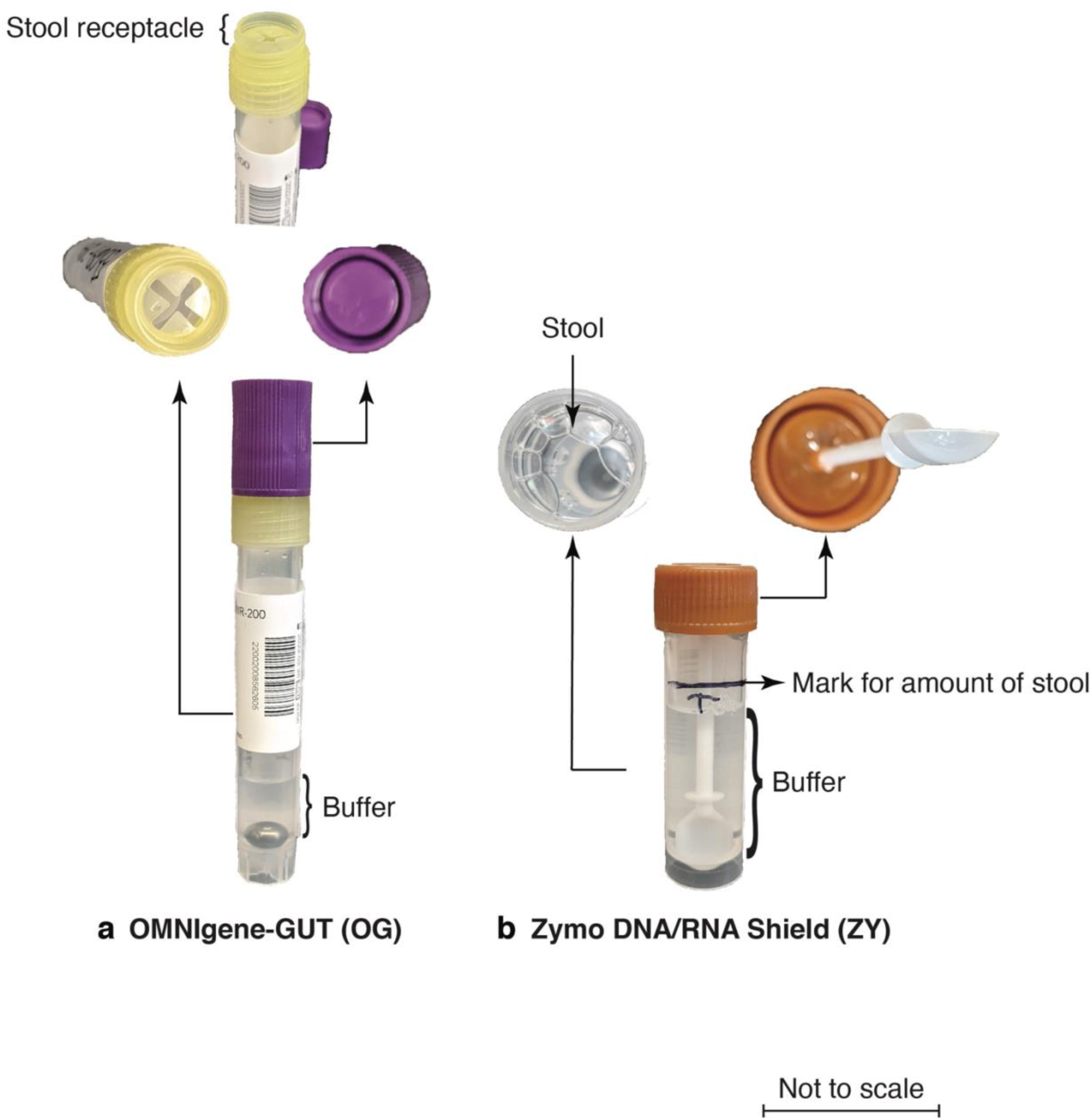
Photographs of the OG and ZY collection and preservation kits. **a.** The OMNIgene-GUT (OG) collection kit includes a special receptacle of defined volume for the collection of stool samples. This is followed by a tube containing 2 ml of proprietary preservative buffer and a metal ball for homogenizing the sample. **b.** Zymo DNA/RNA Shield (ZY) kit is a standard collection tube with the proprietary DNA/RNA shield buffer and plenty of room in the tube above the buffer level for collection of stool.

**Supplementary Fig. 6.**
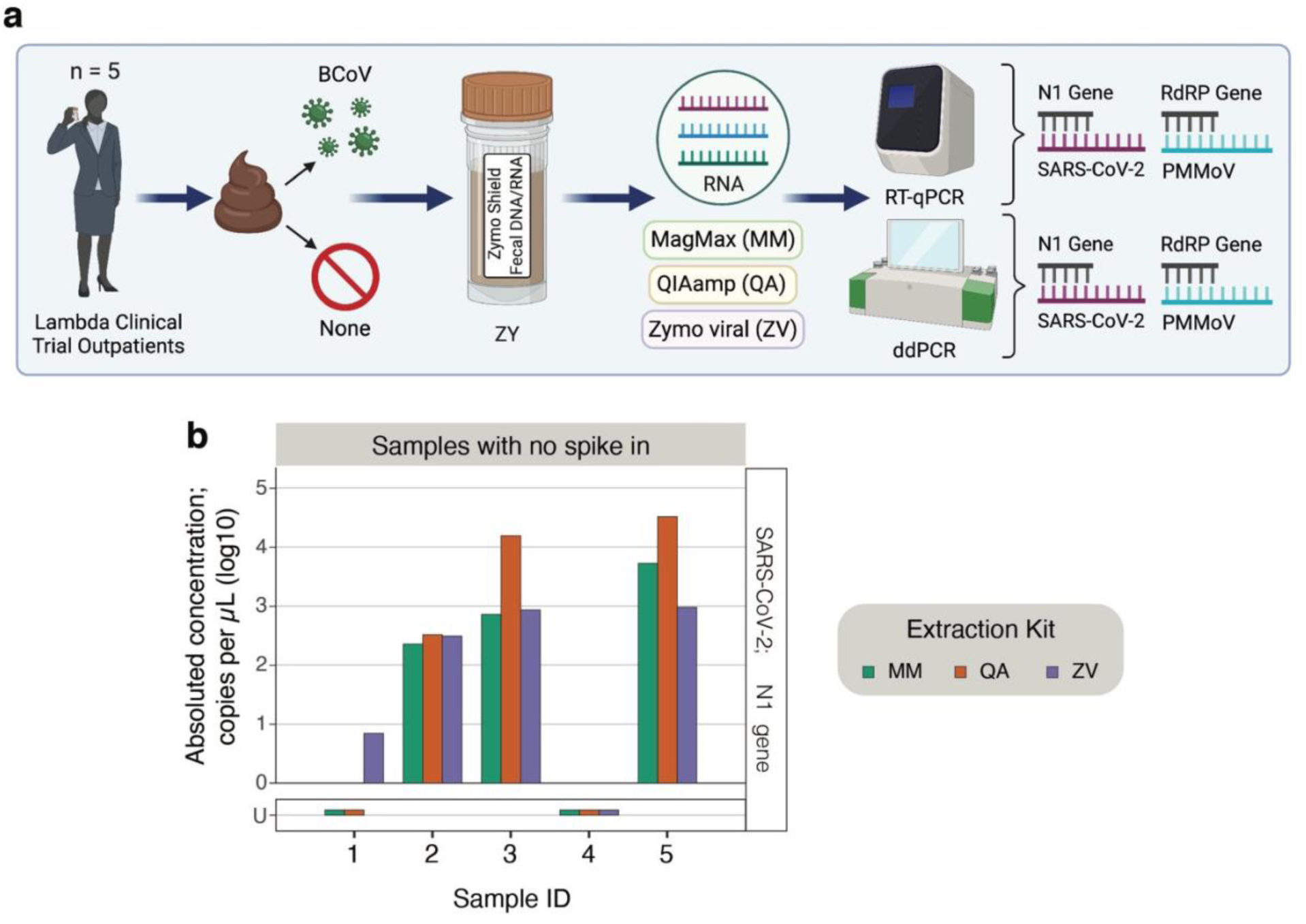
Testing efficiency of three extraction kits using clinical samples stored in the ZY preservative. **a.** Stool samples were collected in the Zymo DNA/RNA shield buffer (ZY) preservative from five COVID-19 outpatients enrolled in a clinical trial of Peginterferon Lambda-1a. All samples were spiked with 10 μl of undiluted BCoV vaccine. In parallel, the same set of samples were also processed without any spike-in. RNA from these samples were extracted using the MagMAX Viral/Pathogen Kit (MM; green), QIAamp Viral RNA Mini Kit (QA; orange) or Zymo Quick-RNA Viral Kit (ZY; purple). **b.** RNA from samples with no spike in were assayed for SARS-CoV-2 RNA using ddPCR targeting the N1 gene. Anonymized sample identities are listed on the x-axis and absolute concentration is listed on the y-axis. ‘U’ stands for undetermined and marks samples with no detectable RNA above LoB. Raw data provided in Supplementary Information 1.

**Supplementary Fig. 7.**
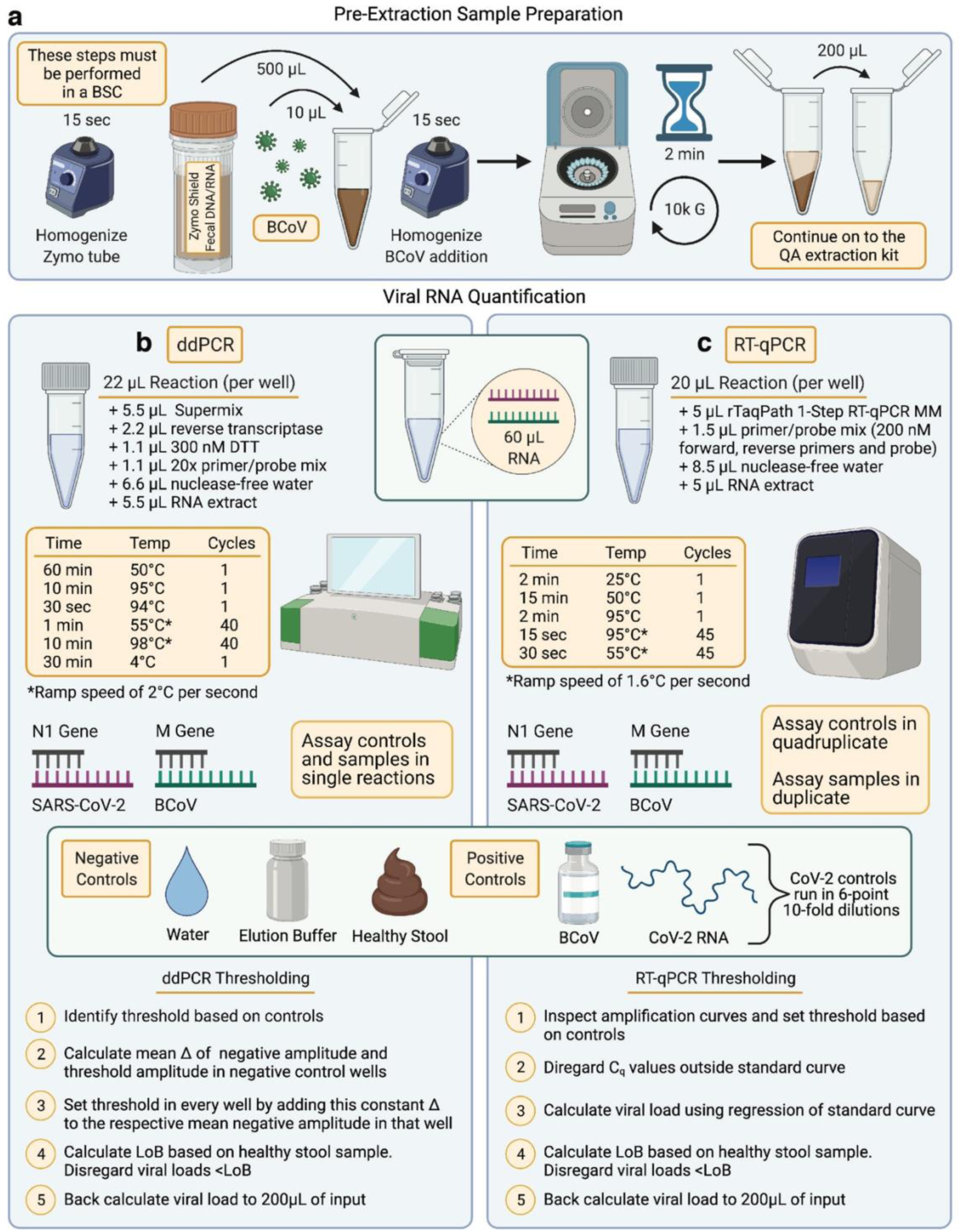
Recommended guidelines for the effective collection and preservation of stool samples for SARS-CoV-2 viral RNA extraction and detection in stool. Illustrated here are recommended guidelines for the detection of SARS-CoV-2 RNA from fecal samples. **a.** Pre-extraction sample preparation. ZY preservation kit is used for the collection of fecal samples from patients. The successive steps are carried out in a biosafety cabinet (BSC). ZY kit with stool is vortexed for 15 seconds. 500 μl of homogenized stool is transferred to an RNAse free, sterile microcentrifuge tube, and spiked with 10 μl of BCoV vaccine. Sample is then vortexed for 15 seconds to ensure uniform mixing of the BCoV control. Spiked in stool aliquot is then centrifuged at 10,000 x g for 2 minutes and 200 μl of the clarified supernatant is transferred to a fresh microcentrifuge tube for RNA extraction using the QA kit as per manufacturer instructions. RNA is eluted in 60 μl of elution buffer. Viral RNA is quantified in this eluate using ddPCR and/or RT-qPCR as follows. **b.** Absolute concentration of RNA is assayed using the one-step RT-ddPCR advanced kit for probes as recommended^19^. Reaction constituents and thermocycling conditions summarized here can be applied to detect the SARS-CoV-2 N1 gene and BCoV M gene. Every assay plate should include three negative controls - water, elution buffer and viral RNA extracted from a healthy stool sample - to determine the Limit of Blank (LoB). Further, every assay plate should include positive controls - QA extracted RNA from BCoV and SARS-CoV-2 synthetic RNA from ATCC. The mean positive and negative amplitudes from these controls are used to guide an appropriate threshold for analysis. The threshold is set between the mean positive and negative amplitudes, such that the negative control does not record presence of viral RNA, while the positive controls reflect the expected RNA concentration. Next, the mean difference between the mean negative amplitude and the threshold amplitude in the negative control reactions is calculated. This difference is added to the mean negative amplitude from every reaction in order to identify a normalized threshold for that specific reaction. **c.** Relative concentration of RNA is assayed using the TaqPath 1-Step RT-qPCR Master Mix, CG as recommended in the manufacturer protocol. Reaction constituents and thermocycling conditions summarized here can be applied to detect the SARS-CoV-2 N1 gene and BCoV M gene. Every 384-well assay plate should include control reactions in quadruplicate. This includes three negative controls - water, elution buffer and viral RNA extracted from a healthy stool sample - to determine the Limit of Blank (LoB). Further, every assay plate should include positive controls - QA extracted RNA from the BCoV vaccine, and a six-point ten-fold dilution series of SARS-CoV-2 synthetic RNA from ATCC starting at 10^4^ copies/μl. Using the control reactions as a reference, inspect the amplification curves of the samples to ensure they are bonafide read-outs and establish a threshold. Disregard Cq values outside the standard curve as “undetermined” since they cannot be used to accurately calculate the viral load. Using a linear regression of the synthetic RNA standards, calculate the relative concentration of viral RNA extracted from the stool sample. Across both ddPCR and RT-qPCR assays, disregard viral loads less than or equivalent to the LoB and back calculate the viral RNA load in the starting samples.

## Supplementary Tables (available as attached files)

**Supplementary Table 1**. Statistical measures from the linear regression of the detection of SARS-CoV-2 viral RNA and BCoV RNA.

**Supplementary Table 2**. Paired t-tests to evaluate the significance of the differential performance of preservatives and extraction kits used with NIST stool samples.

**Supplementary Table 3**. Paired t-tests to evaluate the significance of the differential performance of preservatives and extraction kits used with non-standardized healthy stool samples.

**Supplementary Table 4**. Paired t-tests to evaluate the significance of the differential performance of extraction kits used with clinical samples stored in ZY preservative.

## Notes

### Funding Statement

No authors received payment or services from a third party for any aspect of this submitted work. 
This work was funded by Institutional funds from Stanford University as well as NIH grants that support the Bhatt lab. One of the investigators was also supported by an NSF Graduate Research Fellowship.

### Author Declarations

Stanford University IRB approved this work, as indicated in the main text of the manuscript.

