## Supplementary Information 2 for "Standardized and optimized preservation, extraction and quantification techniques for detection of fecal SARS-CoV-2 RNA"

In this work, we have reported yields of viral RNA across all conditions in the following units: copies of target RNA per μL of stool supernatant. This has been the standard in studies analyzing the shedding of SARS-CoV-2 RNA in stool. However, because a variable amount of stool can be added to the collection kit, the final target RNA concentration is dependent on the amount of stool collected and deposited in each kit. In fact, in a previous analysis, we compare the performance of the OG and ZY preservatives using paired samples from a clinical trial. Here we see that patients tend to provide three-fold more stool in the ZY kit as compared to the OG kit. Concomitantly, we find that samples collected in ZY yield roughly three-times more RNA. Taken together, the amount of stool collected can directly impact the amount of detected target SARS-CoV-2 RNA.

A solution to overcome this challenge of inconsistent loading of stool into a collection tube is to report the yield of SARS-CoV-2 RNA normalized to the weight of input stool. This is the most universal metric and thus allows for comparisons both within and across studies. In the table below, we translate the yields reported from clinical samples preserved in ZY and extracted using all three kits (MM, QA, ZY; Fig. 4) in relation to the amount of stool collected using the following analysis. All variables used herein are described below the equation.

*Copies per g of stool = Copies per µL in eluate * (total eluate from extraction kit / volume of stool supernatant) * (volume of sample / stool weight in sample)*

**Description of variables and related assumptions**

**Copies per µL in eluate:** Absolute concentration derived from ddPCR and normalized by total volume of the reaction (22 μL) and amount of eluate added to the reaction as template (5.5 μL). This assumes a 100% conversion of template RNA to cDNA in the one-step ddPCR reaction. We believe that this conversion is highly efficient, but likely not 100%.

**Total eluate from extraction kit:** Across samples and extraction kits, we eluted the RNA in 60 μL of elution buffer.

**Volume of stool supernatant:** In the ZY kit, we collected stool in the DNA/RNA shield buffer, homogenized the sample by vortexing, and aliquoted 200 μL of the clarified supernatant for extraction after centrifugation. We assume that the virus-like particle and viral RNA are equally distributed across the solid biomass and liquid supernatant of the sample. This is based on preliminary work where resuspension of wastewater samples in the DNA/RNA shield buffer resulted in equivalent distribution across the solid and liquid phases (unpublished data; personal communication from Dr. Marlene Wolfe).

Since the original biobanking effort was not designed to measure weight of stool collected, we sought to find a rational method of calculating the next two variables, post-hoc. Ideally, we know that the ZY collection and preservation kit contains 9 mL of preservative. Further, by measuring the weight of the collection tube before and after collection, we could have been able to calculate the weight of stool deposited. However, in this study we did not measure the weight of the collection tubes before and after collection, and before aliquoting, and thus do not have this information. In future studies, we recommend that these measurements be carried out and noted. In the absence of this information, we set about finding approximations in the following manner. We took two biopsy punches of frozen, homogenized stool and measured the wet weight. Next, we completely dried the stool sample on a heat block set to 100°C for 72 hours and measured the corresponding dry weight. We then assumed that the dried biomass is the amount of stool deposited. The shortcoming of this assumption is that stool is not entirely dry and certainly adds liquid mass to the sample. However, we believe that this assumption will allow us to report concentration per amount of “dry stool” within an order of magnitude from the original “wet stool” concentration. Based on this assumption, we used the dry weight of stool in relation to the total wet weight to calculate the percentage of preserved sample constituted by stool.

**Volume of sample:** 200 μL as used towards viral RNA extraction.

**Stool weight:** Percentage of preserved sample constituted by stool multiplied by 200 μL of sample to calculate the weight of the stool in the volume of sample used for viral RNA extraction.

| **Sample ID** | **Extraction** | **Copies per µL in eluate** | **Percent Stool in Sample by Weight** | **Stool Weight Estimate (g)** | **Copies per g stool** |
| --- | --- | --- | --- | --- | --- |
| 1 | MM | 0 | 0.343 | 0.172 | 0 |
| 1 | QA | 0 | 0.343 | 0.172 | 0 |
| 1 | ZY | 0 | 0.343 | 0.172 | 0 |
| 2 | MM | 27.2 | 0.338 | 0.169 | 24157.89 |
| 2 | QA | 94 | 0.338 | 0.169 | 83486.84 |
| 2 | ZY | 124 | 0.338 | 0.169 | 110131.6 |
| 3 | MM | 152 | 0.325 | 0.163 | 140307.7 |
| 3 | QA | 4928 | 0.325 | 0.163 | 4548923 |
| 3 | ZY | 352 | 0.325 | 0.163 | 324923.1 |
| 4 | MM | 0 | 0.336 | 0.168 | 0 |
| 4 | QA | 0 | 0.336 | 0.168 | 0 |
| 4 | ZY | 0 | 0.336 | 0.168 | 0 |
| 5 | MM | 260 | 0.310 | 0.155 | 251510.2 |
| 5 | QA | 9840 | 0.310 | 0.155 | 9518694 |
| 5 | ZY | 324 | 0.310 | 0.155 | 313420.4 |
